# Genomic and molecular evidence that the lncRNA *DSP-AS1* modulates Desmoplakin expression

**DOI:** 10.1101/2025.03.29.25324867

**Authors:** Luisa Foco, Marzia De Bortoli, M Fabiola Del Greco, Laura S. Frommelt, Chiara Volani, Diana A. Riekschnitz, Benedetta M. Motta, Christian Fuchsberger, Thomas Delerue, Uwe Völker, Tianxiao Huan, Martin Gögele, Juliane Winkelmann, Marcus Dörr, Daniel Levy, Melanie Waldenberger, Alexander Teumer, Peter P. Pramstaller, Alessandra Rossini, Cristian Pattaro

## Abstract

Cardiac desmosomes are specialized cell junctions responsible for cardiomyocytes mechanical coupling. Mutation in desmosomal genes cause autosomal dominant and recessive familial arrhythmogenic cardiomyopathy. Motivated by evidence that Mendelian diseases share genetic architecture with common complex traits, we assessed whether common variants in any desmosomal gene were associated with cardiac conduction traits in the general population.

We analysed data of N=4342 Cooperative Health Research in South Tyrol (CHRIS) study participants. We tested associations between genotype imputed variants covering the five desmosomal genes *DSP, JUP, PKP2, DSG2,* and *DSC2*, and P-wave, PR, QRS, and QT electrocardiographic intervals, using linear mixed models. Functional annotation and interrogation of publicly available genome-wide association study resources implicated potential connection with antisense lncRNAs, DNA methylation sites, and complex traits. Causality was tested via two-sample Mendelian randomization (MR) analysis and validated with functional *in vitro* follow-up in human induced pluripotent stem cell derived cardiomyocytes (hiPSC-CMs).

*DSP* variant rs2744389 was associated with QRS (*P*=3.5×10^-6^), with replication in the Microisolates in South Tyrol (MICROS) study (n=636; *P*=0.010). Observing that rs2744389 was associated with *DSP-AS1* antisense lncRNA but not with *DSP* expression in multiple GTEx-v8 tissues, we conducted two-sample Mendelian randomization analyses that identified causal effects of *DSP-AS1* on *DSP* expression (*P*=6.33×10^-5^; colocalization posterior probability=0.91) and QRS (*P*=0.015). In hiPSC-CMs, *DSP-AS1* expression downregulation through a specific GapmerR matching sequence led to significant *DSP* upregulation at both mRNA and protein levels.

The evidence that *DSP-AS1* has a regulatory role on *DSP* opens the venue for further investigations on *DSP- AS1*’s therapeutic potential for conditions caused by reduced desmoplakin production.

**Author Summary:** Arrhythmogenic Cardiomyopathy is a severe condition mainly caused by pathogenic variants in genes encoding components of the cardiac desmosome, a specialised cell junction.

Given complex traits and Mendelian diseases share common genetic background, we hypothesised that common variants in any of the five desmosomal genes (*DSP, JUP, PKP2, DSG2,* and *DSC2*) could be associated with electrocardiographic measurements in general population individuals.

Analyzing data from >4000 participants from the Cooperative Research In South Tyrol (CHRIS) study, we identified an association between a variant in the desmoplakin gene (*DSP*) and QRS, which represents the time needed for ventricular electrical activation.

Downstream gene expression analyses showed that the identified variant was not associated with the expression of *DSP* but with that of an uncharacterized long non-coding antisense RNA, *DSP-AS1*.

Mendelian randomization (MR) analyses, performed leveraging publicly available data, supported a causal effect of *DSP-AS1* expression on *DSP* expression.

*In vitro* functional follow-up showed that silencing *DSP-AS1* induces *DSP* transcript and desmoplakin protein upregulation, suggesting that *DSP-AS1* is involved in the regulation of *DSP* expression and validating MR findings.

Our study represents a first step in the functional characterization of *DSP-AS1*, a potential target for treatment of diseases caused by low amounts of desmoplakin.

## Introduction

Cardiac desmosomes are specialized cell junctions responsible for cardiomyocytes mechanical coupling. The cardiac desmosome includes the proteins Desmoplakin (*DSP*), Plakophilin-2 (*PKP2*), Desmoglein-2 (*DSG2*), Desmocollin-2 (*DSC2*), and Junction Plakoglobin (*JUP*).

Dysfunctional desmosomes can lead to cardiomyocyte detachment during contraction, altering the mechano-electrical coupling between cells and triggering arrhythmias(1), but also to aberrant activation of signalling pathways such as Wnt/β-catenin signalling(2), the Hippo(3) and TGFβ(4) pathways, which have previously been described to play a pivotal role in ACM pathology, regulating both adipogenesis and fibrogenesis. In fact, pathogenic variants in desmosomal genes have been frequently involved in Arrhythmogenic Cardiomyopathy (ACM), a familial disease, with an autosomal dominant pattern of inheritance with reduced penetrance(5). Homozygous desmosomal gene mutations have also been described to cause recessive forms of ACM(6). ACM is a primary structural cardiomyopathy characterized clinically by life-threatening arrhythmias, increasing the risk of sudden cardiac death. In symptomatic patients, specific electrocardiogram (ECG) abnormalities such as epsilon waves are observed together with palpitations, arrhythmic presyncope/syncope and ventricular tachyarrhythmias(7,8). Despite about 26 genes having been implicated in ACM, the ClinGen Cardiovascular Clinical Domain Working Group has indicated that only the five desmosomal genes *DSP, PKP2, DSG2, DSC2*, and *JUP*, and *TMEM43* are definitively linked to ACM(9).

According to polygenic theory, Mendelian traits can be regarded as extreme manifestations of common complex traits, hence sharing genetic architecture(10). This implies the existence of a spectrum of differential severity observed even within Mendelian phenotypes, indicating that different mutations in the same gene can have a different impact. By extension, a relevant question to ask is whether mutations in ACM genes are associated with altered ECG signatures in individuals not selected for any cardiac disease. For instance, ACM cases were identified in the Finnish general population by typing 6 rare *DSP*, *DSG2*, *DSC2* and *PKP2* variants tested against the PR, QT, QRS, and RR intervals in 6334 individuals, resulting in significant associations with PR at *DSP* and *PKP2*(11).

Expanding this idea, we designed an investigation that assessed whether any common genetic variant located within any of the 5 definitive ACM desmosomal genes (*DSP*, *PKP2*, *DSG2*, *DSC2*, and *JUP*) were associated with cardiac conduction traits in a general population sample. Because ACM patients show conduction abnormalities in depolarization and repolarization, we selected as study outcomes the length of the P-wave, and the PR, QRS, and QT intervals. Building on significant results, we designed Mendelian randomization (MR) experiments to assess if the associated genes had a causal effect on the respective ECG traits. MR was further implemented to assess causal connections between the molecular entities involved by the variant-ECG association, namely mRNA levels of *DSP* and of *DSP* antisense 1 (*DSP-AS1*) long-non-coding RNA (lncRNA), and the cg02643433 methylation. Evidence of a causal effect of the *DSP-AS1* lncRNA on *DSP*, eventually led us to functionally demonstrate the role of *DSP-AS1* on *DSP* gene expression in human induced pluripotent stem cell derived cardiomyocytes (hiPSC-CMs).

## Results

### Genetic association analysis

Discovery and replication study participants characteristics are outlined in **Table 1**. In the CHRIS study, ECG traits were approximately normally distributed, with low-to-null pairwise correlation: the Pearson’s correlation coefficient *r* ranged between 0.09 and 0.21, except for the correlation between PR and P-wave (*r*=0.50; **Supplementary** Figure 1).

**Table 1.**
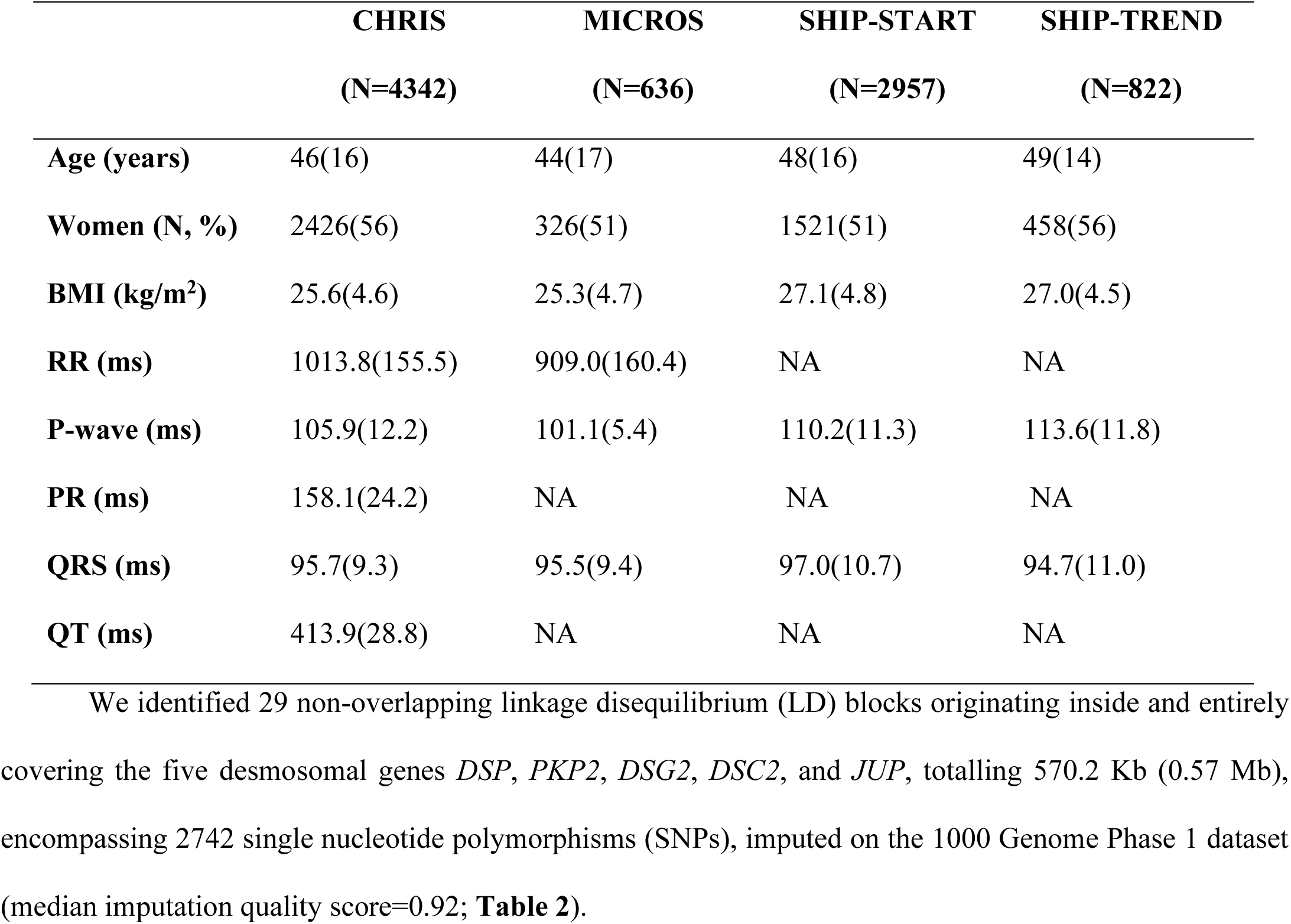
Discovery and replication sample description. ECG statistics are calculated after trait-specific clinical exclusions (**Supplementary** Table 1). Mean and standard deviations (in brackets) describe quantitative variables. Abbreviations: ms, millisecond; NA, not available.

**Table 2.**
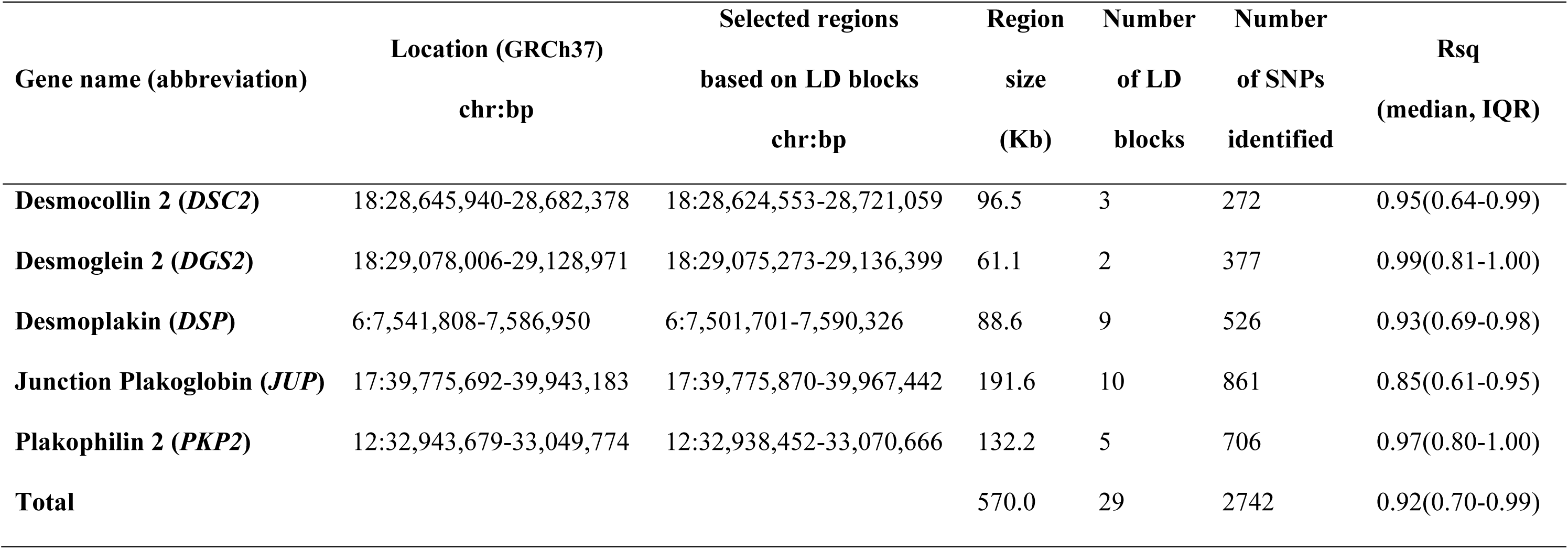
Selected gene regions. Abbreviations: chr, chromosome; bp, base-pairs; LD, Linkage Disequilibrium; Rsq, imputation quality score; IQR, interquartile range.

| Gene name (abbreviation) | Location (GRCh37)<br>chr:bp | Selected regions<br>based on LD blocks<br>chr:bp | Region<br>size<br>(Kb) | Number<br>of LD<br>blocks | Number<br>of SNPs<br>identified | Rsq<br>(median, IQR) |
| --- | --- | --- | --- | --- | --- | --- |
| **Desmocollin 2 (*DSC2*)** | 18:28,645,940-28,682,378 | 18:28,624,553-28,721,059 | 96.5 | 3 | 272 | 0.95(0.64-0.99) |
| **Desmoglein 2 (*DGS2*)** | 18:29,078,006-29,128,971 | 18:29,075,273-29,136,399 | 61.1 | 2 | 377 | 0.99(0.81-1.00) |
| **Desmoplakin (*DSP*)** | 6:7,541,808-7,586,950 | 6:7,501,701-7,590,326 | 88.6 | 9 | 526 | 0.93(0.69-0.98) |
| **Junction Plakoglobin (*JUP*)** | 17:39,775,692-39,943,183 | 17:39,775,870-39,967,442 | 191.6 | 10 | 861 | 0.85(0.61-0.95) |
| **Plakophilin 2 (*PKP2*)** | 12:32,943,679-33,049,774 | 12:32,938,452-33,070,666 | 132.2 | 5 | 706 | 0.97(0.80-1.00) |
| **Total** |  |  | 570.0 | 29 | 2742 | 0.92(0.70-0.99) |

All SNPs were screened for association with the P-wave, PR, QRS, and QT lengths, using EMMAX approximate linear mixed modelling(15), accounting for multiple testing. As displayed by the regional association plots (**Figure 2A**; **Supplementary** Figure 2), we identified significant associations of rs2744389 in *DSP* with QRS (*P*-value=3.7×10^-5^), two nearly independent variants rs115171396 and rs72835665 in *JUP* (LD r^2^=0.017) with P-wave length (*P*-values=2.4×10^-5^ and 6.7×10^-5^), and rs13412, a missense variant in *P3H4* falling within the *JUP* LD region, with QT (*P*-value=1.8×10^-5^; **Supplementary Table 2**). All results were robust to inverse normal transformation of the traits and to adjustment for body mass index (BMI) and the RR interval, except for rs13412, whose association with QT which was not significant anymore after BMI and RR adjustment (**Supplementary Table 2**). We thus excluded rs13412 from further analyses. Significant results were refined using appropriate linear mixed modelling in R (**Table 3**): we observed an effect of -1.10 ms on QRS per copy of the rs2744389 effect allele, which was replicated in the MICROS study (one-sided *P*- value=0.010; **Table 3**), with a very similar effect size of -1.47 ms QRS length per copy of the effect allele. This association did not replicate in the SHIP-START and SHIP-TREND cohorts. The associations of rs115171396 and rs72835665 in *JUP* with P-wave were not replicated in any study (**Table 3**). Analysis of QRS conditional on rs2744389 didn’t identify any additional independently associated variant.

**Figure 1.**
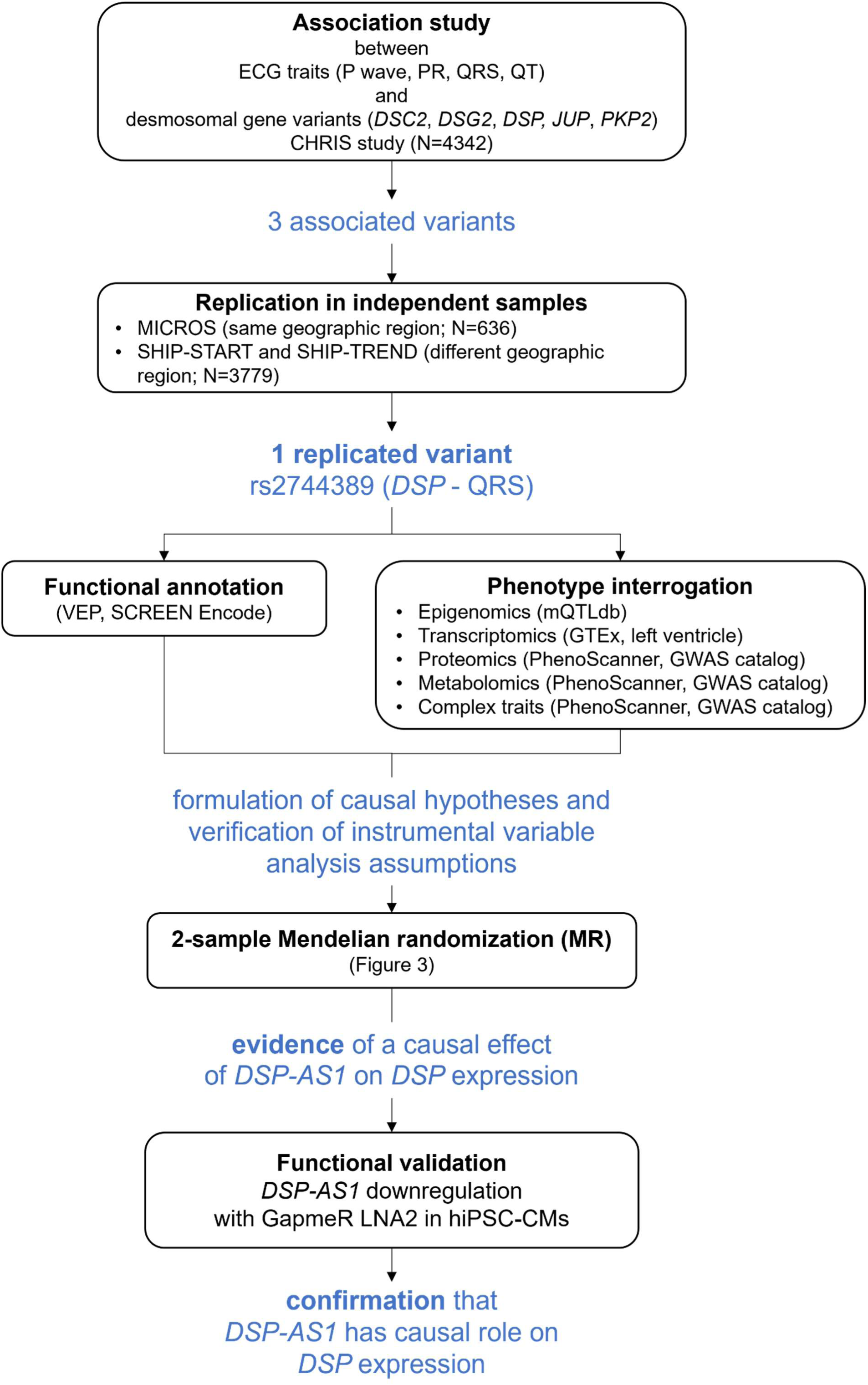
Analysis flowchart and main results.

**Figure 2.**
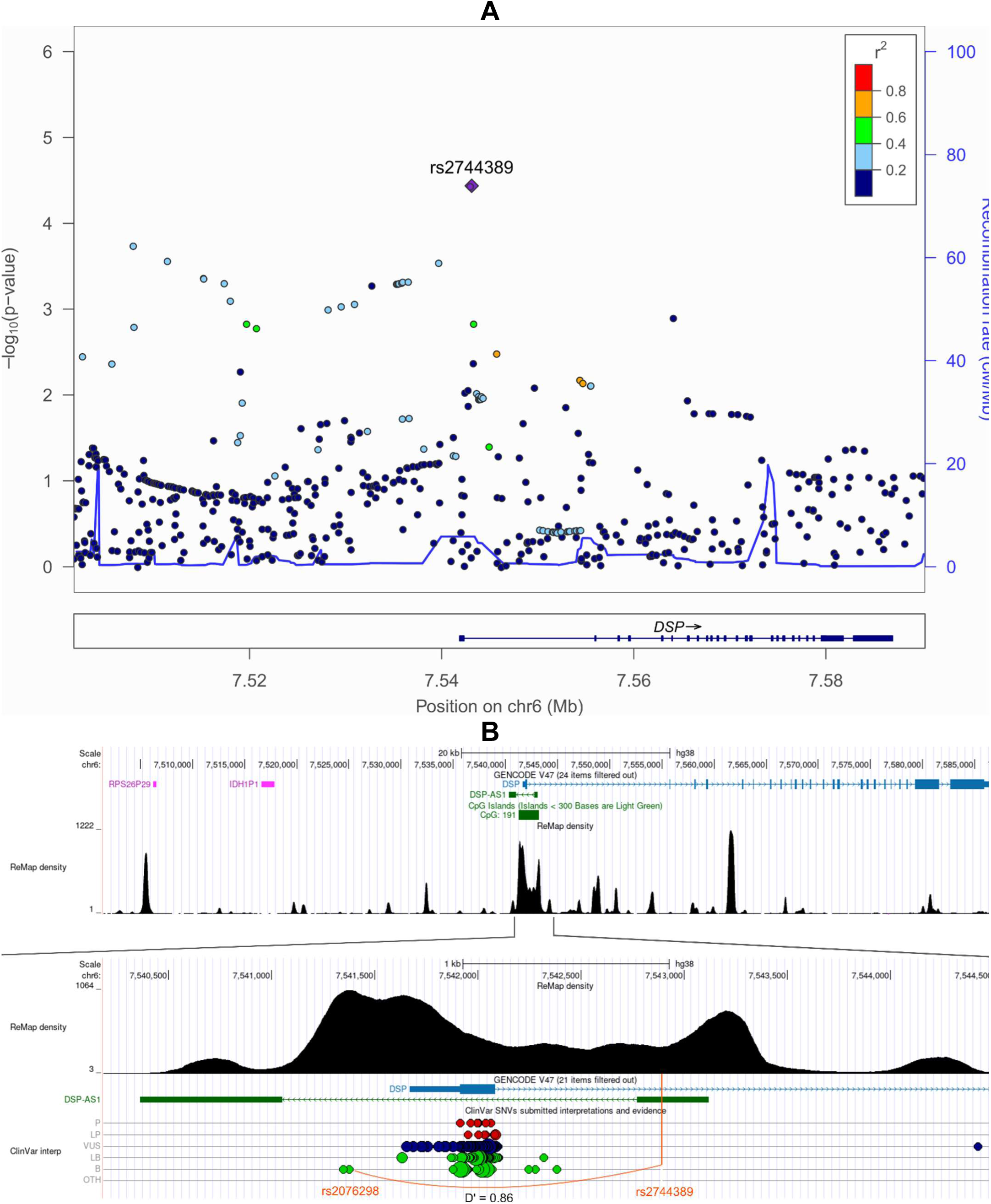
Regional association plot showing association of the *DSP* genomic context with QRS length. Panel. **A**. -log10(*P*-value) of the SNP-QRS association (y-axis) against SNP genomic position (GRCh37; x- axis) at *DSP*. The purple diamond indicates the most associated SNP (position 7,543,123); its LD with the other SNPs is based on the r^2^ estimated on the CHRIS sample. **Panel B**. Annotated genomic context, including validated pseudogenes, ReMap track showing multiple regulatory elements condensed, and ClinVar track. Orange vertical line: rs2744389 location. LD with rs2076298, selected IV for MR, is also highlighted. Figure source: UCSC genome browser.

**Table 3.** Genetic association results. Abbreviations: chr, chromosome; bp, base-pairs; EAF, effect allele frequency; Beta, effect per copy of the effect allele in ms; SE, standard error of Beta. *Build GRCh37 †Reference/Effect allele ‡One-sided.

| Trait | SNP, gene | Chr:bp<sup>*</sup> | Alleles<sup>†</sup> | CHRIS |  |  |  | MICROS |  |  |  | SHIP-START+SHIP-TREND |  |  |  |
| --- | --- | --- | --- | --- | --- | --- | --- | --- | --- | --- | --- | --- | --- | --- | --- |
|  |  |  |  | N | EAF | Beta(SE) | P-value | N | EAF | Beta(SE) | P-value<sup>‡</sup> | N | EAF | Beta(SE) | P-value<sup>‡</sup> |
| P-wave | rs115171396, *JUP* | 17:39910519 | C/T | 4338 | 0.02 | 4.87(1.08) | 6.6×10<sup>-6</sup> | 636 | 0.02 | -0.19(3.12) | 0.525 | 3779 | 0.01 | 1.59(1.45) | 0.136 |
| P-wave | rs72835665, *JUP* | 17:39922558 | G/A | 4338 | 0.51 | -1.10(0.27) | 4.5×10<sup>-5</sup> | 636 | 0.56 | 0.18(0.87) | 0.585 | 3779 | 0.53 | -0.06(0.29) | 0.416 |
| QRS | rs2744389, *DSP* | 6:7543123 | A/C | 4259 | 0.18 | -1.10(0.24) | 3.5×10<sup>-6</sup> | 626 | 0.18 | -1.47(0.64) | 0.010 | 3661 | 0.16 | 0.21(0.32) | 0.748 |

### Functional annotation and phenotypic interrogation

rs2744389 is located in the first intron of *DSP*, immediately downstream the promoter and within a strong enhancer element characterized by H3K4Me1 and H3K27Ac marks, colocalizing with a DNAse I hypersensitivity element; the enhancer is active in the heart right atrium and left ventricle (**Figure 2B**). Additionally, rs2744389 is in linkage disequilibrium with variants found in patients with ACM and cardiocutaneous syndromes (**Supplementary Table 3**). The genomic region selected for association testing included two validated pseudogenes (*RPS26P29* ribosomal protein S26 pseudogene 29, and *IDH1P1* isocitrate dehydrogenase (NADP+) 1 pseudogene 1) and a long non-coding antisense RNA overlapping the DSP promoter, *DSP-AS1* (**Figure 2B**), with rs2744389 falling in the first exon of *DSP-AS1*. The function of the processed pseudogenes is currently unknown, and their expression was nearly undetectable in any tissue of the GTEx v8 dataset (**GTEx Consortium 2020**). *DSP-AS1* has an expression pattern very similar to *DSP*, but ∼10-fold lower (**Supplementary** Figure 3), and its function was unknown.

We interrogated the GWAS catalog and PhenoScanner(16) databases, searching for additional, genome-wide significant associations of rs2744389 with any complex trait, identifying a significant association with pulse rate (*P*-value=1.5×10^-13^) in the UK Biobank(17).

UCSC genome browser interrogation identified SNPs within the analyzed region that were in strong LD with rs2744389 (LD D’>0.80) and significantly associated with ECG traits (**Supplementary** Figure 4): rs7771320 (D’=0.81, r^2^=0.30) with the QRS 12-lead-voltage duration products (12-leadsum)(18); rs112019128 (D’=0.89, r^2^=0.43) with the PR interval(19); rs72825038 (D’=0.94, r^2^=0.46) with PR interval(19) and ECG morphology (amplitude at temporal datapoints(20)); and rs72825047 with spatial QRS-T angle(21). The rs2744389 itself showed some evidence of association with ECG morphology(20) (*P*- value=2×10^-6^).

In GTEx, rs2744389 was associated with *DSP-AS1* expression, which was maximal in the adrenal gland, but not with *DSP* expression (**Supplementary Table 4**). No genome-wide significant DSP eQTL was annotated in GTEx v8. rs2744389 was also associated with methylation of cg02643433, located in the CpG island 201 in the first intron of *DSP*: this association was consistent across multiple datasets (**Supplementary Table 4**). We didn’t observe evidence of associations with protein levels or metabolites.

### Mendelian randomization (MR) and colocalization analyses

Motivated by this overall evidence, rather than focusing on the functional characterization of the intronic rs2744389, which looks more a classical “tag” variant, we hypothesized that cg02643433 and *DSP-AS1* could regulate *DSP* expression, which could in turn causally affect QRS duration. We therefore tested the concatenation of causal effects depicted in **Figure 3** using the two-sample MR technique. We first tested the causal effect of cg02643433 methylation on the RNA levels of *DSP-AS1* (**Figure 3, pathway A**) and *DSP* (**Figure 3, pathway B**), and on QRS duration (**Figure 3, pathway C**). To this end, we extracted genetic variants associated with cg02643433 methylation from mQTLdb(22), choosing methylation levels from blood of the middle age group, and from GTEx v8 heart left ventricle, which was the most relevant tissue available for the current investigation.

**Figure 3.**
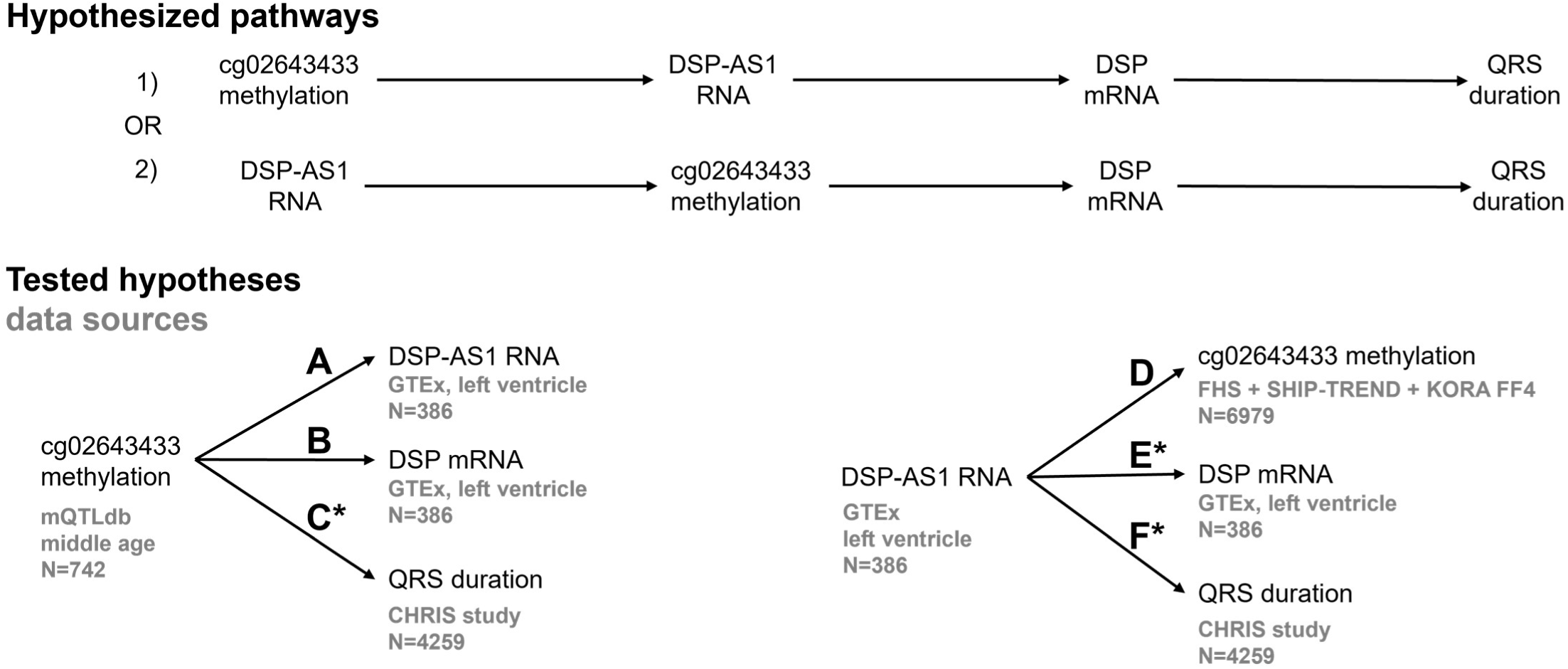
Mendelian randomization analysis scheme. Upper panel: overview of the two hypothesized biological pathways underlying *DSP* regulation, possibly contributing to QRS duration. Lower panel: decomposition of the pathways by individual analysis with indication of the instrumental variable and data sources. Causal effect of cg02643433 methylation on *DSP-AS1* mRNA level (**A**), *DSP* mRNA level (**B**), and QRS duration (**C**); causal effect of *DSP-AS1* RNA level on cg02643433 methylation (**D**), *DSP* mRNA (**E**, experimentally validated), and QRS duration (**F**). *DSP* mRNA and desmoplakin protein levels were excluded due to lack of genome-wide significant associations. *denotes significant MR results (**Table 4**).

**Table 4.**
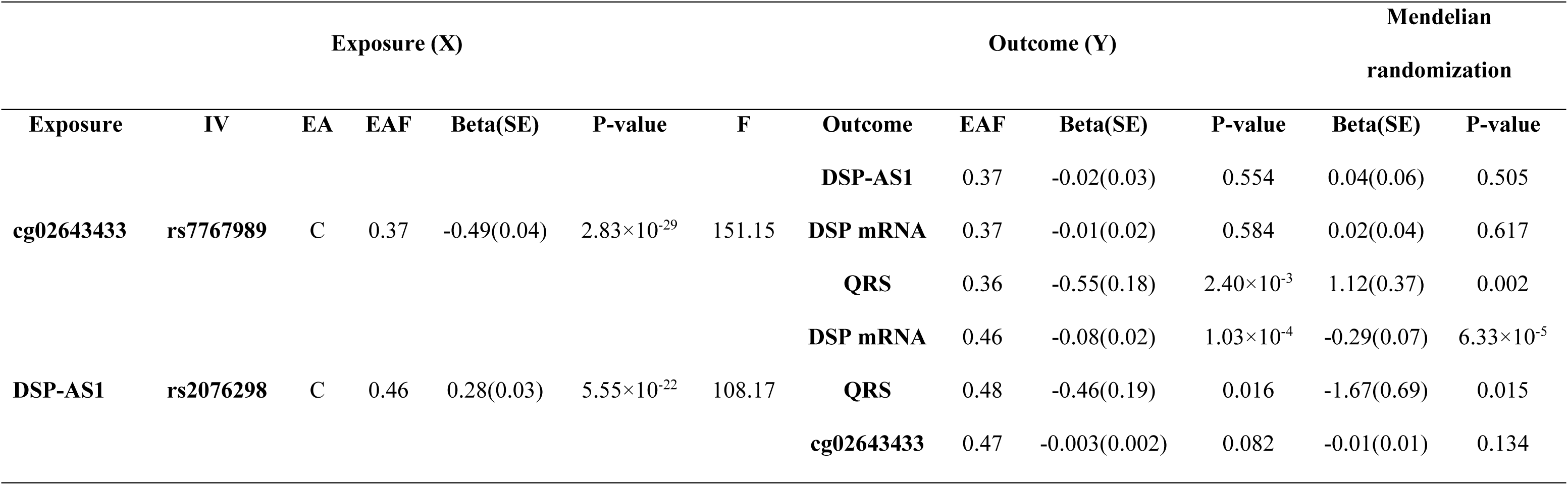
Mendelian randomization analysis results. Results of the analyses presented in **Figure 3**. Exposure summary statistics obtained from: mQTLdb database, middle age N=742 (cg02643433); GTEx, left ventricle N=386 (*DSP-AS1*). Outcome summary statistics obtained from: GTEx left ventricle N=386 (*DSP-AS1; DSP*); CHRIS study N=4259 (QRS); FHS N=4170, SHIP-TREND N=964, KORA FF4 N=1928 (cg02643433). EA, Effect Allele; EAF, Effect Allele Frequency. Beta, effect per copy of the effect allele; SE, standard error of Beta.

Next, we tested the causal effect of *DSP-AS1* expression on *DSP* expression (**Figure 3, pathway D**), on QRS duration (**Figure 3, pathway E**) and reverse causation on cg02643433 methylation (**Figure 3, pathway F**). The latter was tested in consideration of biological evidence showing that antisense RNA can modify DNA methylation(23). We extracted genetic variants associated with *DSP-AS1* expression from the GTEx v8 left ventricle dataset and from the Framingham Heart Study (FHS)(24), the SHIP-TREND and KORA FF4 studies in whole blood for testing reverse causation (**Figure 3, pathway F**).

Because we could not find any SNP genome-wide significantly associated with *DSP* mRNA level in the left ventricle, and no desmoplakin protein GWAS was available, we couldn’t test further questions such as for instance whether the desmoplakin protein levels affect QRS duration.

Studies used to identify the IV summary statistics are described in **Supplementary Table 5**. As strong IVs (*P*-value<5×10^-8^; F statistic>10), we identified 26 and 119 SNPs associated with cg02643433 methylation (**Supplementary Table 6**) and *DSP-AS1* expression in left ventricle (**Supplementary Table 7**), respectively. LD pruning (r^2^ >0.01; **Supplementary Datasets 13 and 14**) left a single IV per exposure: rs7767989 for cg02643433 and rs2076298 for *DSP-AS1* (**Figure 2B**, **Table 4**). Scientific literature examination showed that selected IVs were not associated with other traits than the selected exposures. Variants in LD with them were associated with cardiovascular and ECG-related phenotypes or pulmonary phenotypes. The rs2076295 was associated with levels of the receptor for advanced glycation product RAGE, a cell surface pattern receptor recognizing multiple ligands, mostly expressed in the lung and involved in inflammation. Overall, available evidence suggests that vertical pleiotropy could exist (cardiovascular traits) but horizontal pleiotropy violating MR assumptions is unlikely (**Supplementary Table 8**).

cg02643433 methylation was causally associated with increased QRS duration (*P*-value=0.002); *DSP- AS1* expression was causally associated with decreased QRS duration (*P*-value=0.015; **Table 4**) and decreased *DSP* expression in the heart left ventricle (*P*-value=6.33×10^-5^). This latter finding supports an antisense-mediated mechanism of gene expression regulation, where an antisense RNA (*DSP-AS1*) downregulates its target (*DSP*). Confounding by LD was ruled out by evidence of colocalization between *DSP-AS1* and *DSP* expression in left ventricle (PPH4=0.91; **Supplementary Table 9**). Colocalization between *DSP-AS1* expression and QRS could not be proven (PPH4=0.09), likely because the QRS association peak was not pronounced (PPH1=0.73).

### *DSP-AS1* and *DSP* mRNA expression in different cell types

To validate the causal effect of *DSP-AS1* on *DSP* expression (**Figure 3 pathway E**), we conducted a series of *in vitro* experiments. We first tested whether *DSP-AS1* and *DSP* mRNA were expressed in keratinocytes, human induced pluripotent stem cells (hiPSCs), hiPSC-derived cardiomyocytes (hiPSC-CMs), and HEK cells, using digital droplet PCR (ddPCR). Both transcripts were expressed in all tested cells, at various degrees, with keratinocytes and HEK cells expressing the highest and lowest *DSP* levels, respectively. *DSP-AS1* was less expressed than *DSP* in all cell lines, showing the highest expression in HEK and the lowest in hiPSC-CMs (**Supplementary** Figure 6). We therefore proceeded with the functional follow-up, downregulating *DSP-AS1* in hiPSC-CMs.

### *DSP-AS1* downregulation in hiPSC-CMs

We treated hiPSC-CMs with two GapmeRs, LNA1 and LNA2, plus a negative control GapmerR, LNA_NC, testing different conditions. All GapmeRs were delivered to cells by gymnosis. The LNA2 induced *DSP-AS1* downregulation under all tested conditions. LNA1 treatment showed no effect, like LNA_NC, for which treated cells didn’t show differences compared to non-treated cells (NT) (**Supplementary** Figure 7). We repeated hiPSC-CMs treatment with LNA2 at 1000nM for 10 days, confirming a ∼2.5-fold downregulation of *DSP-AS1* compared to LNA_NC-treated cells (**Figure 4A**).

**Figure 4.**
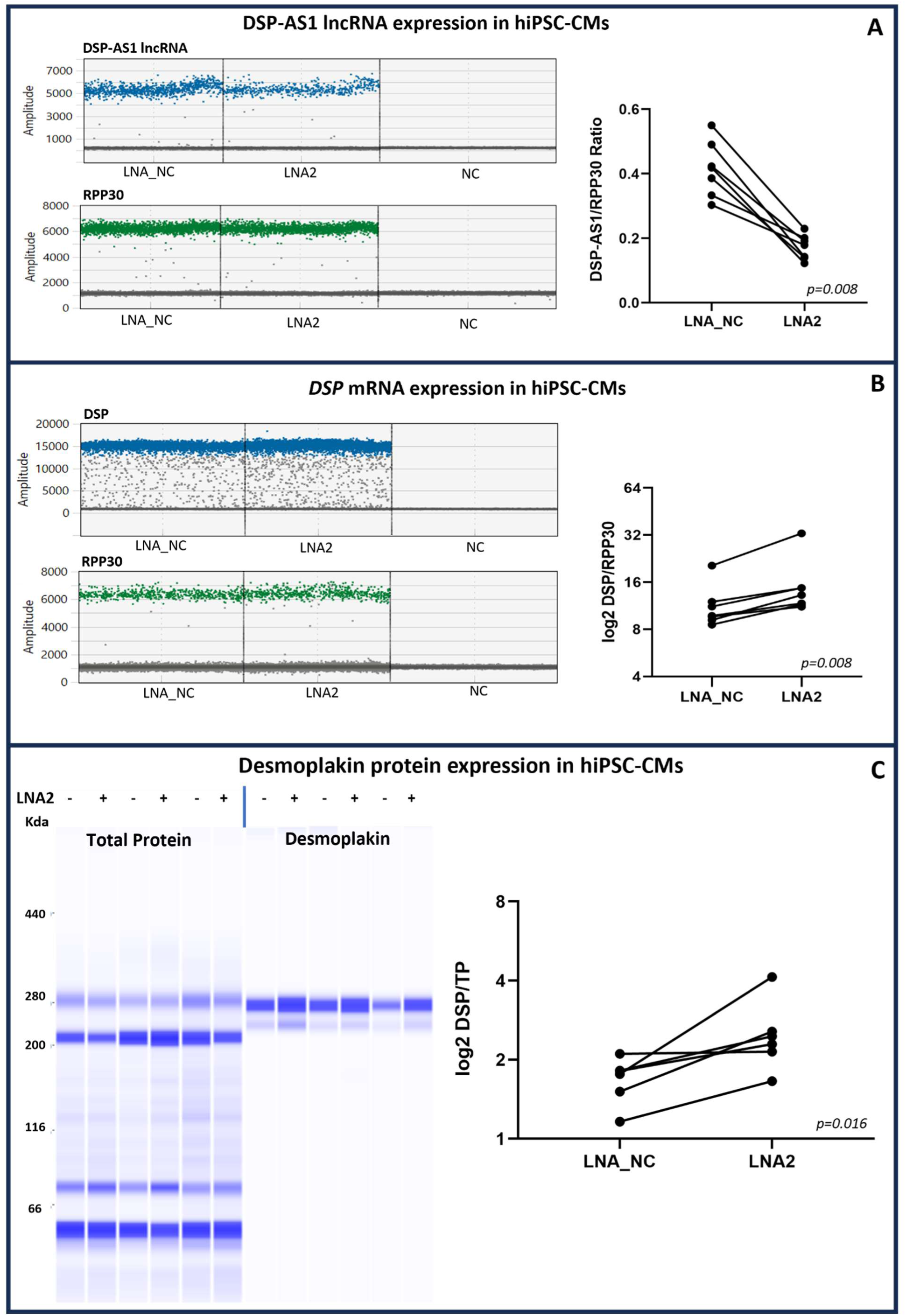
*In vitro* validation of the causal effect of *DSP-AS1* on *DSP* mRNA and protein levels. Effect of treatment of hiPSC-CMs with LNA control (LNA-NC) and LNA2 GapmerR at 1000nM for 10 days (LNA2). Data were available on 7 independent cardiomyogenic differentiations. Each dot in the pairplot represents an independent differentiation; lines in the pairplot connect the same differentiation to highlight the change in the measured outcomes after treatment with LNA2. **A)** Relative expression of *DSP-AS1* after LNA2 treatment (*P*-value=0.008). **B)** Relative expression of DSP mRNA after LNA2 treatment (*P*- value=0.008). NC: negative control; RPP30: reference gene; amplitude indicates the fluorescence intensity of each probe; in blue, the FAM channel for *DSP-AS1* and *DSP;* in green, the HEX channel for RPP30. **C)** Relative abundance of desmoplakin protein after LNA2 treatment (*P*-value=0.016). TP: Total protein. All data were analysed using a one-sided Wilcoxon matched-pairs signed rank test. Raw data are provided in **Supplementary Tables 13, 14 and 15**.

### *DSP-AS1* downregulation leads to an increased Desmoplakin mRNA and protein levels in hiPSC-CMs

We then tested whether *DSP-AS1* downregulation could affect *DSP* mRNA expression using ddPCR, observing evidence of a significant ∼1.5-fold increase in *DSP* mRNA level following *DSP-AS1* downregulation by LNA2 (**Figure 4B**). We finally tested whether desmoplakin protein levels were also affected by *DSP-AS1* downregulation using the Protein Simple Wes™ system, observing a protein upregulation of ∼1.5-fold in LNA2-treated cells (**Figure 4C**).

## Discussion

By combining genomic and clinical data from population-based studies with *in vitro* functional experiments, we demonstrated that downregulating *DSP-AS1* lncRNA expression causes an increase of *DSP* mRNA and protein level. Additionally, *DSP-AS1* resulted being causally associated with QRS duration.

LncRNAs have been recently recognized as key regulators of multiple cellular functions, from the arrangement of chromatin architecture to the regulation of RNA transcription and post-transcriptional modifications, protein synthesis and localization(23). However, they remain poorly characterized by scarce functional studies. In cardiovascular disease, dysregulated lncRNAs contribute to various mechanisms, including endothelial dysfunction and myocardial remodeling and some of them serve as biomarkers for disease diagnosis and prognosis(25).

Previous studies identified three *DSP-*targeting lncRNAs in non-cardiac tissues and cell lines: *MIR4435-2HG*(26) and *UPLA1* (*GJD3-AS1*)(27), respectively promoting gastric cancer and lung adenocarcinoma progression, that target desmoplakin, leading to Wnt/β-catenin signaling pathway activation; and *LYPLAL1-AS1*(28), which directly binds desmoplakin, possibly targeting it to proteasome degradation, resulting in Wnt/β-catenin pathway signaling downregulation and human adipose-derived mesenchymal stem cells adipogenic differentiation. In summary, lncRNA-mediated *DSP* downregulation leads to both Wnt/β- catenin pathway activation (cancer cells) and inhibition (adipogenic differentiation), a contrasting behaviour that has been previously reported in ACM(29).

Our findings add a novel, *cis*-acting mechanism of antisense-mediated *DSP* gene regulation, differently from the above mentioned *trans*-acting lncRNAs. *DSP* and *DSP-AS1* are transcribed bidirectionally, with divergent transcription, overlapping with a head-to-head configuration(30). Hypermethylation upstream and immediately downstream of the *DSP* transcriptional start site overlapping *DSP-AS1* was detected in lung cancer cell lines, leading to decreased *DSP* levels and Wnt/β-catenin pathway activation as previously reported(31). Differential methylation of the *DSP* conserved promoter was also observed in human and mouse lung epithelial cells seeded on matrigel-coated soft or stiff polyacrylamide gels, showing opposite effects(32). We speculate that *DSP-AS1* may be involved in the regulation of *DSP* expression controlling chromatin architecture acting on DNA methylation, a hypothesis that we have tested but could not be confirmed within our MR framework (p=0.134). However, as our MR analyses were based on summary statistics available on methylation data from blood only, and DNA methylation is tissue specific, additional experiments are warranted on cardiac-specific methylation data.

Clarifying *DSP-AS1* mechanism of action is relevant, also in light that ACM caused by *DSP* pathogenic variants is currently regarded as a distinct clinical entity called Desmoplakin cardiomyopathy. Patients exhibit acute myocardial injury episodes, inflammation and extensive left ventricular involvement even at early disease stages, with an aggressive arrhythmic course, that was recently characterized in depth(33,34). A review of the ARVD/C Genetic Variants Database (https://arvc.molgeniscloud.org/menu/main/home) revealed seven variants in exon 1 (p.Gln51X; p.Val30Met; p.Tyr42=; p.Gly35=; p.Gly46Asp; p.Leu26=; p.Thr49Ser) and one variant in 5’UTR (c.1dupA). It is therefore crucial to ascertain the impact of variants in the *DSP-AS1* regions that overlap with *DSP*, to establish a more precise genotype-phenotype relation. Loss of functional *DSP* was rescued in a zebrafish model, through genetic and pharmacological manipulations, leading to Wnt/β- catenin activation and consequent beneficial effects(35). Findings disagree with a more recent work showing that suppression rather than activation of the Wnt/β-catenin pathway is beneficial in desmoplakin cardiomyopathy(36). Altogether, *DSP* appears as an actionable target that could be manipulated by acting on *DSP-AS1*, resulting in a possible strategy for increasing desmoplakin protein level in *DSP* haploinsufficient ACM patients. Because *DSP* is also involved in dilated cardiomyopathy, cardio cutaneous disorders, cancer and lung diseases, targeting *DSP-AS1* could also have additional applications.

All causal and downstream analyses started by observing the QRS-rs2744389 association in CHRIS, which was replicated in MICROS, a small study conducted in the same Alpine area(13). MICROS participants who joined the CHRIS study were removed from the analysis, guaranteeing separate samples. The similarity of the allelic effects on QRS between the two studies is remarkable. The association did not replicate in SHIP-START and SHIP-TREND, conducted in Northern Germany. The reason is unclear and could lie in the characteristics of the locus, perhaps sensitive to environmental exposures or particularly structurally unstable. Despite all participants to the four studies were of European descent, there might be differences in the genetic architecture in terms of population-specific LD patterns(37), which was observed to affect genomic associations also within Europe(38). Local adaptation seems less plausible. CHRIS and MICROS participants are living in a mountain region at moderate-to-high altitude as opposed to SHIP participants living at the sea level. ECG is known to change in response to chronic altitude exposure(39), but the variability of the genetic association would imply a gene-environment interaction that is unproven. As also the CHARGE consortium GWAS of QRS(40) did not report genome-wide significant associations at this locus, it is likely that we are in the presence of either genomic instability(41) or environmental interaction. Nevertheless, genome-wide significant associations with QRS-related traits were observed for SNPs in LD with rs2744389: rs7771320 (D’=0.81) associated with the QRS complex 12-lead sum(18) and rs72825038 (D’=0.94) associated with QRS morphology(20). In LD were also rs112019128 and rs72825038, previously associated with PR interval by the CHARGE consortium(19) but not significant in CHRIS. These findings corroborate the relevance of *DSP* and its promoter region in association with ECG regulations in general population individuals, not necessarily affected by cardiac pathologies.

Our work has both strengths and limitations. We depicted a pipeline for a candidate gene approach and downstream analyses, applying MR to prioritize potential causal targets for subsequent functional follow-up. The main strength is the *in vitro* validation of the causal effect of *DSP-AS1* on *DSP* gene observed with MR. The validation is particularly relevant because it overcomes a major limitation of our two-sample MR analysis between *DSP-AS1* and *DSP* expression, where the IV summary statistics for both exposure and outcome were extracted from the same GTEx heart left ventricle sample, resulting in full overlap. Sample overlap may bias MR estimates(42), even if a recent investigation showed reassuring results that two-sample MR methods can be applied to a one-sample framework without major risks of bias(43). Another major limitation was the impossibility to test all possible causal links in the depicted pathways, due to the absence of appropriate IVs for some variables. Additionally, methylation QTL were derived from blood and not from cardiac tissues. We were also unable to perform a multivariate MR, due to presence of a single IV for the exposures of interest.

## Conclusions

In conclusion, genomics analyses of desmosomal genes in association with ECG traits in a general population sample, identified a variant associated with QRS length located in the promoter region of *DSP*, at the *DSP-AS1* lncRNA. Downstream causal analyses provided evidence of a potential novel antisense-mediated mechanisms controlling *DSP* expression in *cis*. This evidence was finally corroborated by *in vitro* GapmeR analysis, proving that *DSP-AS1* can regulate *DSP* expression both at mRNA and protein levels. Additional experimental investigations are warranted to clarify the *DSP-AS1* mechanisms of action. These should include a fine analysis of the *DSP-AS1/DSP* promoter sequence to determine the impact of variants on *DSP* expression and on the three-dimensional structure, stability and function of *DSP-AS1*. While *DSP-AS1* represents a potential target for treatment of diseases caused by *DSP* mutations, such as ACM, dilated cardiomyopathy, cardiocutaneous diseases (e.g. Carvajal syndrome) and some cancer conditions, further investigations are warranted to test the identified GapmeR on hiPSC-CMs carrying a desmoplakin mutation causing a protein deficit. Such models could ideally envision the use of advanced engineered heart tissue platforms, efficiently simulating physiological conditions(44).

## Materials and methods

### Study design

Considered in these analyses were 4342 participants to the Cooperative Health Research in South Tyrol (CHRIS) study, with complete genotype and electrocardiographic data included in the second CHRIS data release on participants recruited between 2011 and 2014. Briefly, the CHRIS study is a population-based cohort study being conducted since 2011 in the Val Venosta/Vinschgau district (South Tyrol, Italy)(12,45). Data include socio-demographic, health, and lifestyle information collected through questionnaires-based interviews and quantitative traits assessed through clinical examinations and urine and blood sampling under overnight fasting conditions.

Replication of genetic associations was tested in MICROS and SHIP.

The Microisolates in South Tyrol Study (MICROS)(13) was a cross-sectional, population-based study conducted in three Alpine villages of the same Val Venosta/Vinschgau district where also the discovery CHRIS study was conducted. Considered for replication were 636 individuals who did not participate to the CHRIS study, to guarantee sample independence.

The Study of Health in Pomerania (SHIP-TREND) is a longitudinal population-based cohort study in West Pomerania, a region in the northeast of Germany, assessing the prevalence and incidence of common population-relevant diseases and their risk factors. Baseline examinations for SHIP-TREND were carried out between 2008 and 2012, comprising 4,420 participants aged 20 to 81 years. Study design and sampling methods were previously described (**Völzke 2022**). The CHRIS analyzed data can be requested for research purposes to the CHRIS Access Committee at, and via https://transfer.ship-med.uni-greifswald.de for SHIP.

### Ethics approval and consent to participate

CHRIS was approved by the Ethics Committee of the Healthcare System of the Autonomous Province of Bolzano (approval number 21-2011). MICROS was approved by the Ethics Committee of the Autonomous Province of Bolzano on 26-February-2002 (approval number RD/ac/13/01/18332). The research involving human stem cell lines was approved by the Ethics Committee of the Province of Bolzano (approval number 1/2014). SHIP-START and SHIP-TREND were approved by the Ethics Committee at the University Medicine Greifswald (approval number BB 39/08). The studies conform to the Declaration of Helsinki, and with national and institutional legal and ethical requirements. All participants included in the analysis gave oral and written informed consent.

### Study outcomes and exclusions

Primary outcomes were the duration of the P-wave, PR, QRS and QT intervals, reflecting atrial and ventricular depolarization and repolarization. We obtained data from 10 seconds ECGs performed using standard 12-lead ECG workstations: PC-ECG-System Custo 200 – Customed (CHRIS); Mortara Portrait, Mortara Inc., Milwaukee, USA (MICROS); Personal 120LD, Esaote, Genova, Italy (SHIP-START and SHIP-TREND). In all studies, participants were asked to remain silent and in supine position during the procedure. Participants with history of atrial fibrillation, myocardial infarction, heart failure, Wolff-Parkinson-White syndrome, assuming class I and III antiarrhythmics and/or digoxin, pacemaker carriers, and pregnant women, were excluded from the analyses as detailed in **Supplementary Table 1**. Values of ECG traits outside the range (first quartile – 3*interquartile range) and (third quartile + 3*interquartile range) were further removed. Between-trait pairwise correlations were estimated by the Pearson’s correlation coefficient.

### Genotyping and selection of the genomic intervals

CHRIS and MICROS DNA samples were genotyped using the Illumina HumanOmniExpressExome Bead array. Genotyped SNPs were retained if they had call rate >99%, Hardy Weinberg Equilibrium (HWE) *P*- value≥3×10^-8^, and minor allele frequency ≥0.01. Samples with evidence of sex mismatch, duplication and labelled as outliers after principal component analysis were removed. Data were imputed against the 1000 Genome Phase 1 dataset using ShapeIT2 and Minimac3(46), on GRCh37 assembly.

In SHIP-START, DNA samples were genotyped on the Affymetrix Genome-Wide Human SNP Array 6.0. Excluded were samples with call rate <86% and SNPs with position mapping issues, HWE *P*- value≤0.0001, call rate ≤0.8, or monomorphic. In SHIP-TREND, DNA samples were genotyped on the Illumina Human Omni 2.5 array. Excluded were samples with call rate <94% and SNPs with position mapping issues, HWE *P*-value≤0.0001, call rate≤0.9 or monomorphic. Samples were excluded from both SHIP-START and SHIP-TREND in case of duplication or sex mismatch. Both studies imputed their genotypes with IMPUTE v2.2.2(47) against 1000 Genomes Phase I (interim).

Genotyping of KORA S4/F4 samples was performed on the Affymetrix Axiom Platform. Genotypes were called with the Affymetrix software and annotated to NCBI build 37. Genotype data quality control steps included for the samples, a call rate of 97%, mismatch of phenotypic and genetic gender, 5 standard deviations from mean heterozygosity rate, check for European ancestry, check for population outlier, comparison with other genotyping of the same individuals (Metabochip, Exome, Omni). For the SNPs, a call rate of 98% was used, an adherence p-value to Hardy-Weinberg equilibrium of 5×10^-10^ and an MAF<0.02. Then, imputation was performed based on the 1000G Phase 3 reference panel, using minimac3 as imputation tool (imputation done on Michigan Imputation server) and with SHAPEIT v2 as a pre-phasing tool.

### Methylation data SHIP

DNA was extracted from blood samples of n=508 SHIP-TREND participants to assess DNA methylation using the Illumina HumanMethylationEPIC BeadChip array. Samples were randomly selected based on availability of multiple OMICS data, excluding type II diabetes, and enriched for prevalent MI. The samples were taken between 07:00 AM and 04:00 PM, and serum aliquots were prepared for immediate analysis and for storage at -80 °C in the Integrated Research Biobank (Liconic, Liechtenstein). Processing of the DNA samples was performed at the Helmholtz Zentrum München. Preparation and normalization of the array data was performed according to the CPACOR workflow(48) using the software package R (www.r-project.org). The array idat files were processed using the minfi package. Probes that had a detection p-value above background (sum of per-array methylated and unmethylated intensity values based p-value ≥1E-16) were set to missing.

Methylation beta values were calculated as proportion of methylated intensity value on the sum of methylated+unmethylated+100 intensities. Arrays with observed technical problems (±4SD outside control probe intensity mean) during steps like bisulfite conversion, hybridization or extension, as well as arrays with mismatch between sex of the proband and sex determined by the chr X and Y probe intensities were removed from subsequent analyses. Additionally, only arrays with a call rate ≥ 95% were processed further resulting in 495 samples with methylation data on 865,859 sites available for subsequent analyses.

DNA methylation of additional 480 samples of the SHIP-TREND baseline cohort was assessed using the HumanMethylationEPICv2 BeadChip array and processed using the same workflow as before, of which 476 samples passed final quality control. For this array type, the call rate of the final samples was >85%.

Technical replicate probes were merged using the *sesame* R package.

### Methylation data KORA FF4

The KORA (Kooperative Gesundheitsforschung in der Region Augsburg) research platform has been collecting clinical and genetic data from the general population in the region of Augsburg, Germany for over 20 years. The KORA FF4 (2013–2014) cohort (n=2,279) is a follow-up study from the KORA S4 (n=4,261) survey carried out 1999-2000. In the baseline examinations all inhabitants of German nationality between the ages of 25 and 74 years were enrolled. Participants completed a lifestyle questionnaire, including details on health status and medication use, underwent standardized examinations with blood samples taken(49).

Genomic DNA from 1928 individuals was bisulfite converted using the EZ-96 DNA Methylation Kit (Zymo Research, Orange, CA, USA) in two separate batches (N=488, N=1440). Subsequent methylation analysis was performed on an Illumina (San Diego, CA, USA) iScan platform using the Infinium MethylationEPIC BeadChip according to standard protocols provided by Illumina. GenomeStudio software version 2011.1 with Methylation Module version 1.9.0 was used for initial quality control of assay performance and for generation of methylation data export files. Further quality control and preprocessing of the data were performed in R v3.5.1 (R Core Team (2017). R: A language and environment for statistical computing. R Foundation for Statistical Computing, Vienna, Austria. URL https://www.R-project.org/), with the package minfi v1.28.3(50) and following primarily the CPACOR pipeline(48). Raw intensities were read into R (command read.metharray) and background corrected (bgcorrect.illumina). Probes with detection p-values >0.01 were set to missing.

Before normalization, we removed problematic samples and probes. Forty samples were removed: 2 showed a mismatch between reported sex and that predicted by minfi; 33 had median intensity <50% of the experiment-wide mean, or <2000 arbitrary units; and 9 (overlap of 4 with previous) had >5% missing values on the autosomes. A total of 59631 probes were removed (some overlapping multiple categories): cross-reactive probes as given in published lists (N=44493; (51,52)); probes with SNPs with minor allele frequency >5% at the CG position (N=11370) or the single base extension (N=5597) as given by minfi; and 5786 with >5% missing values. A total of 806228 probes remained for analysis. Quantile normalization (QN) was then performed separately on the signal intensities divided into the 6 probe types: type II red, type II green, type I green unmethylated, type I green methylated, type I red unmethylated, type I red methylated(48). For the autosomes, QN was performed for all samples together; for the X and Y chromosomes, men and women were processed separately. The transformed intensities were then used to generate methylation beta values, a measure from 0 to 1 indicating the percentage of cells methylated at a given locus.

Probes from the X chromosome (N=17743, following quality control) and the Y chromosome (N=379) were excluded from the analysis.

### Genetic association analysis in the CHRIS study

In the CHRIS genomic dataset, we selected the regions encompassing linkage disequilibrium (LD) blocks originating inside the desmosomal genes *DSP*, *PKP2*, *DGS2*, *DSC2*, and *JUP*, and extending outside the gene boundaries. LD-blocks were defined based on the D’ confidence intervals(53) and identified applying LDExplorer to the 1000 Genome phase 3 European-ancestry panel(54).

Association between dosage levels and ECG traits was tested using a genome-wide association study- like approach based on EMMAX as implemented in EPACTS v3.2.6(15), adjusting for age and sex, assuming a genetic additive model, and accounting for relatedness, estimated on the genotyped autosomal variants. The statistical significance level was set at 2.6×10^-4^, corresponding to the ratio between the genome-wide significance level of 5×10^-8^ to the fraction of genome tested (the LD regions around the 5 desmosomal genes covered approximately 3000 megabases; **Table 2**). Significantly associated variants were re-tested using appropriate linear mixed model fitting through *lmekin* function implemented in the R package ‘coxme’ v2.2.5: models included fixed effects for age and sex, and random effect for the day of recruitment, to remove potential long-term recruitment effects(45). Relatedness was modeled as in EMMAX within the variance-covariance matrix.

We performed two sensitivity analyses: additionally adjusting for RR interval and BMI; and applying the rank-based inverse normal transformation to the ECG traits. Regional association plots were generated with LocusZoom v0.4.8(55). Full results of EMMAX analyses and scripts are provided in **Supplementary Datasets 1-12 and Supplementary Appendix**.

### Replication testing

Direction-consistent replication was tested, based on the same genetic model, in the MICROS study by fitting linear mixed models adjusted for age, sex, village, and relatedness, using the *lmekin* function as above, and in SHIP-START and SHIP-TREND by fitting a simple linear models adjusted for age and sex. The Bonferroni-correct significance level for replication was set to 0.017 (0.05 over 3 variants tested for replication).

### metQTL Analysis SHIP

To account for potential confounding effects due to blood cell composition, blood cell subtypes were measured and included in the association model. Additionally, the array processing batch (n=248 and n=247) for the EPIC arrays, and the first six principal components of the control probe intensities obtained by the CPACOR workflow were included in the model to account for technical factors. The EPIC and EPICv2 arrays were analyzed separately. Details on assessment of the metabolic phenotypes and covariates used in this analysis are provided within the SHIP cohort design. The association analyses were conducted in R.

### metQTL Analysis KORA

First, beta values were transformed using the inverse-normal transformation. This was followed by calculation of methylation residuals, wherein a linear regression model was used with adjustment for: sex, age, smoking, technical covariates, white blood cell counts.

As technical variables, the principal components of the (non-negative) control probes were determined and the top 20 were used as covariates in the models, as per the CPACOR pipeline. A “batch” variable (a 0/1 covariate) was also adjusted for, since the KORA FF4 methylation data consists of two measurement rounds. The white blood cell counts used are monocytes, basophils, eosinophils, and neutrophils, they were measured in the KORA FF4 cohort.

Second, we performed the metQTL analysis, using the R package MatrixEQTL. The linear regression model implemented was DNA methylation residual ∼ SNP, considering only cis-associations with a cis distance of 1 Mbp. We used dosages of the alternate allele.

Additionally to the general QC of the KORA F4 genotype data, we further excluded variants with MAF<0.05. For the genetic variants, the ID chr:positions_REF_ALT was used, with REF/ALT=reference/alternate allele according to the reference panel 1000G.

### rs2076298 meta-analysis

Prior to MR analysis, we performed a fixed-effects meta-analysis of the association between rs2076298 and cg02643433 in FHS, KORA FF4 and SHIP-TREND using the command *metan* implemented in Stata 18. Pooled estimate was used for subsequent analyses and are shown in **Supplementary** Figure 5.

### Variant annotation

Associated SNPs were annotated using the Ensembl Variant Effect Predictor tool available in Ensembl GRCh37 (http://www.ensembl.org/Tools/VEP), the UCSC genome browser (**genome.ucsc.edu**), and the SCREEN Encode tool (https://screen.encodeproject.org/) (GRCh37). LD of the CHRIS sample was estimated using LocusZoom v0.4.8(55), using the –vcf option.

### Phenotype interrogation

We checked whether the associated variants were also associated with other traits at the genome-wide significance level of 5×10^-8^, including diseases, DNA methylation levels, gene expression, and protein levels, interrogating the PhenoScanner v2 (last accessed on 13/02/2024)(16), the Genotype-Tissue Expression GTEx Project database v8(56), and the methylation mQTLdb database(22).

### Mendelian randomization (MR) analyses

We conducted two-sample MR analyses selecting SNPs as instrumental variables (IV) and retrieving summary statistics of association from published GWAS(22,40,56) and from the current analysis in CHRIS. To satisfy the assumption of relevance, we selected genetic variants associated with the exposure at genome-wide significance level (p<5×10^-8^) with F statistic >10. To ensure IVs independency, we selected variants with LD r^2^ <0.01. LD was estimated using the Ensemble LD Calculator using the 1000 Genome phase 3 European ancestry panel as reference (https://www.ensembl.org/Homo_sapiens/Tools/LD).

For each exposure, we could identify only one single SNP satisfying the MR assumptions for use as IV. To exclude the presence of pleiotropy, and hence verifying the exclusion restriction assumption, we inspected available biological evidence from the literature. Following effect allele and direction harmonization between exposures and outcomes, MR estimates were computed as the Wald ratio estimate, with the standard error derived via delta method approximation, using the R package ‘MendelianRandomization’ v0.9.0(57). Because we were interested in dissecting two alternative pathways, significance level was set at 0.05/2=0.025. MR scripts and data are provided in **Supplementary Appendix** and **Supplementary Dataset 15.**

### Colocalization analyses

We performed statistical colocalization analysis(58) of *DSP-AS1* expression in left ventricle with *DSP* expression in the same tissue and with QRS duration (**Figure 3, pathways E, F**), using the *coloc* package v 5.1.0 implemented in R. The data used for the analyses described in this manuscript were obtained from the GTEx Portal on 11/15/24. All analyses were conducted within ±100 kb from rs2076298, which was selected as instrumental variable in the MR analysis.

### Functional follow-up

#### Cell cultures

For the initial analysis of endogenous *DSP* and lncRNA expression, we used human induced pluripotent stem cells (hiPSC), hiPSC-derived cardiomyocytes (iPSC-CMs), commercial adult human primary keratinocytes (HPK), and human embryonic kidney HEK293T cell lines. HPK were cultured in keratinocyte growth medium (Human EpiVita Serum-Free Growth Medium 141-500a, Cell Applications). HEK293T cells were grown as previously described(59). The hiPSCs line available for this study derives from one healthy individual who was previously characterized(60,61). Briefly, hiPSCs were cultured in feeder-free conditions on 6-well plates coated with Matrigel (Corning), using a ready-to-use, commercially available medium (StemMACS™ iPS- Brew XF; Miltenyi Biotec). The cardiomyogenic differentiation was induced with the PSC Cardiomyocyte Differentiation Kit (Thermo Fisher Scientific). After 22-25 days of cardiomyogenic differentiation, the beating monolayer of cells was dissociated by Multi Tissue Dissociation Kit 3 (Miltenyi Biotec) to obtain a purified iPSC-CMs population, through the depletion of non-CMs cells, by using PSC-Derived Cardiomyocyte Isolation Kit (Miltenyi Biotec). The purified hiPSC-CMs were replated on matrigel coated 24-well plates (150.000-200.000 cells/well) in a basal medium (High Glucose DMEM; Gibco), 2% Hyclone Fetal Bovine Defined (GE Healthcare Life Sciences), 1% non-essential Amino Acids (Gibco), 1% Penicillin/Streptomycin (Gibco) and 0.09% β-mercapto-ethanol (Gibco) for further experiments. After 3-4 days of recovery, the purified hiPSC-CMs were treated with GapmerRs as described below.

#### GapmeRs design and delivery in hiPSC -CMs

For the lncRNA knockdown, specific locked nucleic acid (LNA) antisense GapmeRs targeting RP3-512B11.3 lncRNA (Transcript Annotation ENST00000561592.1_1) were designed using the Qiagen RNA Silencing tool, available at https://geneglobe.qiagen.com/us/customize/rna-silencing. The tool ranked several GapmeRs at the highest score. Two were selected, named LNA1 and LNA2, and tested in hiPSC-CMs together with the GapmerR negative control A (NC; LG00000002-FFA, Qiagen).

Four different conditions were tested, at GapmeRs concentrations of: 1) 500nM for 5 days; 2) 500nM for 10 days; 3) 1000nM for 5 days; and 4) 1000nM for 10 days. In all conditions, the delivery of GapmeRs into hiPSC-CMs was performed through an unassisted “naked” uptake, that is, GapmeRs were directly added to cell medium without transfection reagents. This approach, also known as *gymnosis*, is less toxic for the cells and shows a higher efficiency in cells that are difficult to be transfected as hiPSC-CMs(62,63). To perform *gymnosis*, the *in vivo ready* high-quality grade (HPLC purification with a final step of sodium salt exchange) was required for the GapmeRs production. For conditions 2 and 4, after 5 days of treatment, the culture medium was replaced with fresh medium containing the same concentration of GapmerR.

#### RNA isolation and ddPCR analysis

Total RNA was extracted from cultured cells by TRIzol reagent and Direct-zol RNA Kit (Zymo Research). Then, reverse transcription of 50ng RNA was performed using SuperScript VILO cDNA Synthesis Kit (Invitrogen) in a total volume of 20 µl. Droplet Digital PCR (ddPCR) was performed using a QX200 system (Bio-Rad) according to manufacturer’s recommendations. The reactions (22μl total volume) contained 2× ddPCR™ Supermix for Probes (no dUTP) (Bio-Rad), 20× primer/probe assay for each target, except for *DSP* for which it contained 31.5× primer/probe assay, 1 ng of cDNA for *DSP* and 4ng of cDNA for the *DSP-AS1* lncRNA, and water up to the total volume. For the specific detection of the lncRNA *DSP-AS1*, *DSP* and the reference *RPP30* gene, the following primer/probe assays were used: Bio-Rad qhsaLEP0147498 (FAM), Bio-Rad dHsaCPE5047954 (FAM) and Bio-Rad dHsaCPE5038241 (HEX), respectively. The droplets were generated with the QX200™ Droplet Generator (Bio-Rad), mixing 20 μl of the reactions described above and 70 μl of Droplet Generation Oil for Probes (Bio-Rad), loaded in the proper lanes of DG8™ cartridges. Droplets were then transferred to a 96-well PCR semi-skirted plate and the reaction was performed using a GeneAmp™

PCR System 9700 (Applied Biosystems), according to the following program: 95 °C for 10 min, then 45/40 (*DSP*/*DSP-AS1*) cycles of (94 °C for 30 s, 57/60 °C (*DSP*/*DSP-AS1*) for 2 min), 98 °C for 10 min and 4 °C for the storage. Amplification signals were read using the QX200™ Droplet Reader (Bio-Rad) and analyzed using the QuantaSoft software (Bio-Rad). All ddPCR details are described following the Minimum Information for Publication of Digital PCR Experiments (dMIQE) guidelines checklist(64) and are available in **Supplementary Table 10**.

#### Desmoplakin protein expression analysis

RIPA lysis buffer, composed of 10 mM Tris-HCl pH 7.4, 150 mM NaCl, 1% Igepal CA630 (NP-40), 1% sodium deoxycholate (NaDoc), 0.1% SDS (Sodium Dodecyl Sulfate), 1% Glycerol, supplemented with protease and phosphatase inhibitors (Complete Tablets, Mini EASYpack, Roche) was used to lysate hiPSC- CMs, after GapmeRs treatment. The protein level was quantified using Pierce™ BCA Protein Assay Kit (Thermo Scientific).

hiPSC-CMs lysates were tested for desmoplakin protein (sc-390975 mouse anti desmoplakin I/II (A- 1), Santa Cruz) and total protein (Total Protein Detection Module, Bio-Techne) using a 66–440 kDa Separation Module (Bio-Techne) on the Protein Simple Wes™ system (Bio-Techne). Lysates were diluted with 0.1X Sample Buffer to a final concentration of 0.2 μg/μl, then mixed with 5X Fluorescent Master Mix and heated at 95 °C for 5 min. Mouse anti-desmoplakin antibody was used at 1:25 dilution, total protein biotin labelling reagent reconstitution mix was prepared following the manufacturer’s instructions and then loaded with prepared samples and other reagents (ladder, blocking antibody diluent, HRP-conjugated anti mouse secondary antibody (Anti-Mouse Detection Modules, Bio-Techne), total protein streptavidin HRP and the luminol-peroxide mixture) in the assay plate. We used the following specific instrument settings: *total protein size* as assay type, separation run at 475 V for 30 min, incubation time of 30 min for total protein biotin labelling, total protein streptavidin HRP, primary and secondary antibodies. High dynamic range (HDR) function was applied for luminol/peroxide chemiluminescence detection.

#### Data analysis

Results of these laboratory experiments were visually inspected via paired dot-plots (“pairplot”). Given hiPSC-CMs obtained from the same differentiation were split into two groups, one treated with LNA2 and the other with LNA-CN, distributions of mRNA expression and protein levels were compared across using the Wilcoxon matched-pairs signed rank test to account for the matched conditions. Because prior evidence was either available or could be hypothesized for the direction of the effect, we applied a one-sided test.

Statistical analyses of data generated from the laboratory experiments were performed using GraphPad version 9.3.1 (471). All experimental raw data are available in **Supplementary Tables 11-15**.

## Data Availability

The CHRIS analyzed data can be requested for research purposes to the CHRIS Access Committee at, and via https://transfer.ship-med.uni-greifswald.de for SHIP.

## Acknowledgements

We thank all CHRIS, MICROS, SHIP, KORA, and FHS participants.

Full acknowledgements for the CHRIS study are reported at http://translational-medicine.biomedcentral.com/articles/10.1186/s12967-015-0704-9#Declarations. CHRIS biobank “Bioresource Research Impact Factor” (BRIF) code: BRIF6107.

The SHIP authors are grateful to Paul S. DeVries for his support with the EWAS pipeline.

We thank all participants for their long-term commitment to the KORA study, the staff for data collection and research data management and the members of the KORA Study Group (https://www.helmholtz-munich.de/en/epi/cohort/kora) who are responsible for the design and conduct of the study.

We thank all collaborators of both Eurac Research and the Healthcare System of the Autonomous Province of Bolzano who made the CHRIS and MICROS studies possible. We thank Emilio Cusanelli for his precious suggestions on lncRNA’s science. We thank the Department of Innovation, Research, University and Museums of the Autonomous Province of Bolzano-South Tyrol for covering the Open Access publication costs.

## Supporting information caption

S1 Table. Exclusion criteria applied to each ECG trait.

S2 Table. Results of EMMAX main and sensitivity analyses.

S3 Table. rs2744389 Linkage Disequilibrium with DSP-AS1 and DSP variants in ClinVar.

S4 Table. Additional traits associated with rs2744389.

S5 Table. Mendelian Randomization study details.

S6 Table. SNPs associated with cg02643433 in middle age at genome-wide significance level extracted from the mQTLdb (Gaunt 2016).

S7 Table. SNPs associated with DSP-AS1 at genome-wide significance level extracted from GTEx left ventricle (v8).

S8 Table. Traits associated at genome-wide significance level in the DSP region.

S9 Table. Colocalization results.

S10 Table. The Minimum Information for Publication of Digital PCR Experiments (dMIQE) guidelines checklist.

S11 Table. Raw data from ddPCR quantification of DSP-AS1 and DSP mRNA in different cell types.

S12 Table. Raw data from GapmeR experimental setting.

S13 Table. Raw data from GapmeR DSP-AS1 downregulation.

S14 Table. Raw data from GapmeR DSP downregulation.

S15 Table. Raw data from desmoplakin protein after DSP-AS1 GapmeR downregulation.

S1 Figure. Distributions of the ECG traits in the CHRIS study sample S2 Figure. Regional association plot for P-wave over the JUP region. S3 Figure. DSP and DSP-AS1 expression profile in multiple tissues.

S4 Figure. Linkage disequilibrium of rs2744389 with all nearby variants associated with ECG traits at P<5×10-8.

S5 Figure. Fixed effects meta-analysis of association between rs2076298 and cg02643433.

S6 Figure. DSP-AS1 and DSP mRNA expression in different cell types.

S7 Figure. GapmeRs set up for DSP-AS1 downregulation.

S1 Dataset. Summary statistics of association of variants in desmosomal genes with P wave.

S2 Dataset. Summary statistics of association of variants in desmosomal genes with PR. S3 Dataset. Summary statistics of association of variants in desmosomal genes with QRS. S4 Dataset. Summary statistics of association of variants in desmosomal genes with QT.

S5 Dataset. Summary statistics of association of variants in desmosomal genes with P wave – sensitivity.

S6 Dataset. Summary statistics of association of variants in desmosomal genes with PR – sensitivity. S7 Dataset. Summary statistics of association of variants in desmosomal genes with QRS – sensitivity. S8 Dataset. Summary statistics of association of variants in desmosomal genes with QT – sensitivity.

S9 Dataset. Summary statistics of association of variants in desmosomal genes with inverse normal P wave – sensitivity.

S10 Dataset. Summary statistics of association of variants in desmosomal genes with inverse normal PR – sensitivity.

S11 Dataset. Summary statistics of association of variants in desmosomal genes with inverse normal QRS – sensitivity.

S12 Dataset. Summary statistics of association of variants in desmosomal genes with inverse normal QT – sensitivity.

S13 Dataset. LD in the region including variants associated with cg02643433 at GWAS level.

S14 Dataset. LD in the region including variants associated with DSP-AS1 at GWAS level.

S15 Dataset. Raw data used to conduct MR analysis.

Supplementary Appendix. Scripts used for data analysis.

---

**Supplementary Figure 1.**
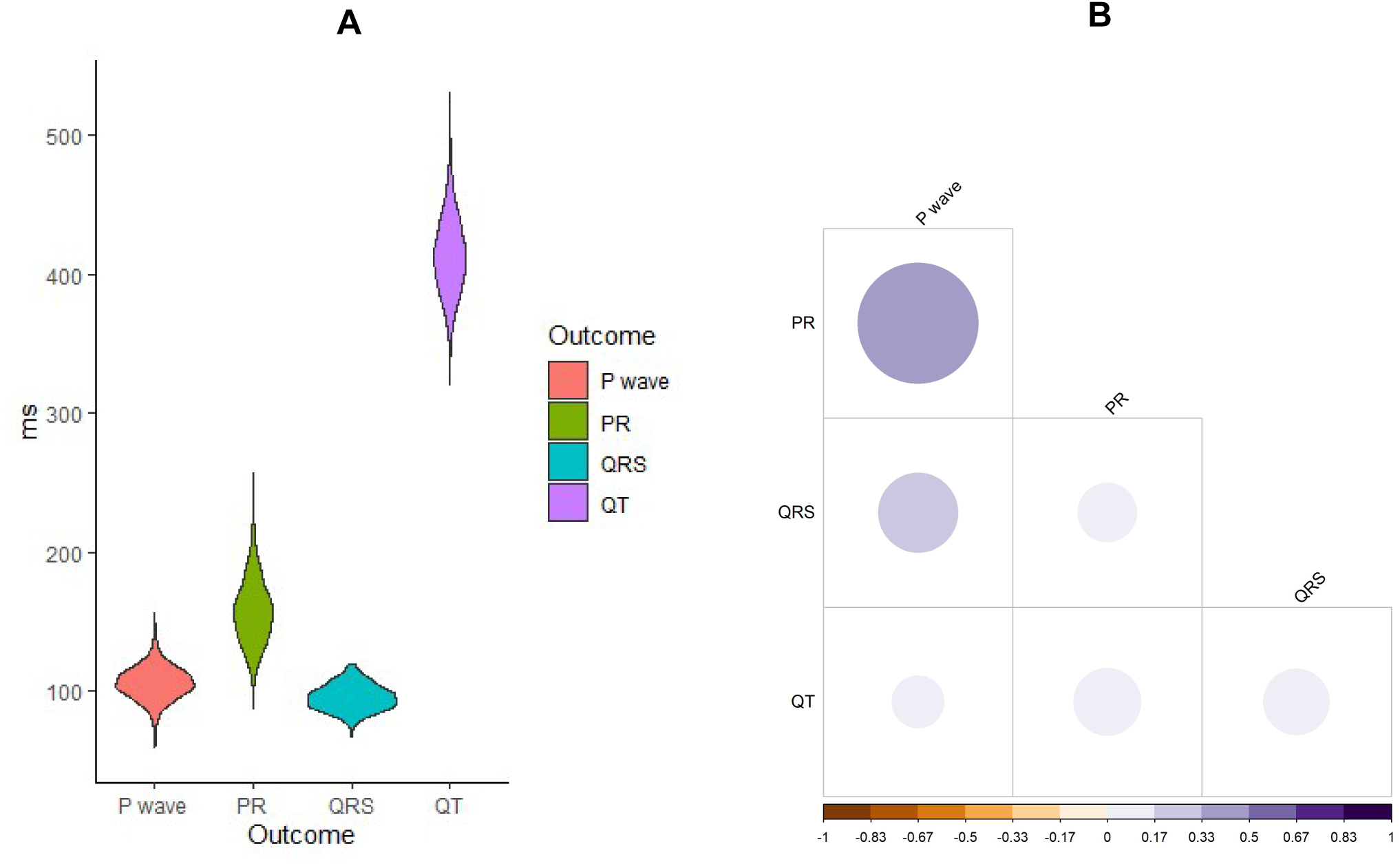
Distributions of the ECG traits in the CHRIS study sample. Panel. **A**. Violin plots of P-wave, PR, QRS, and QT, showing their distributions in milliseconds (ms; y-axis). **Panel B**. Heatmap of the pairwise Pearson’s correlation coefficients among all ECG traits.

**Supplementary Figure 2.**
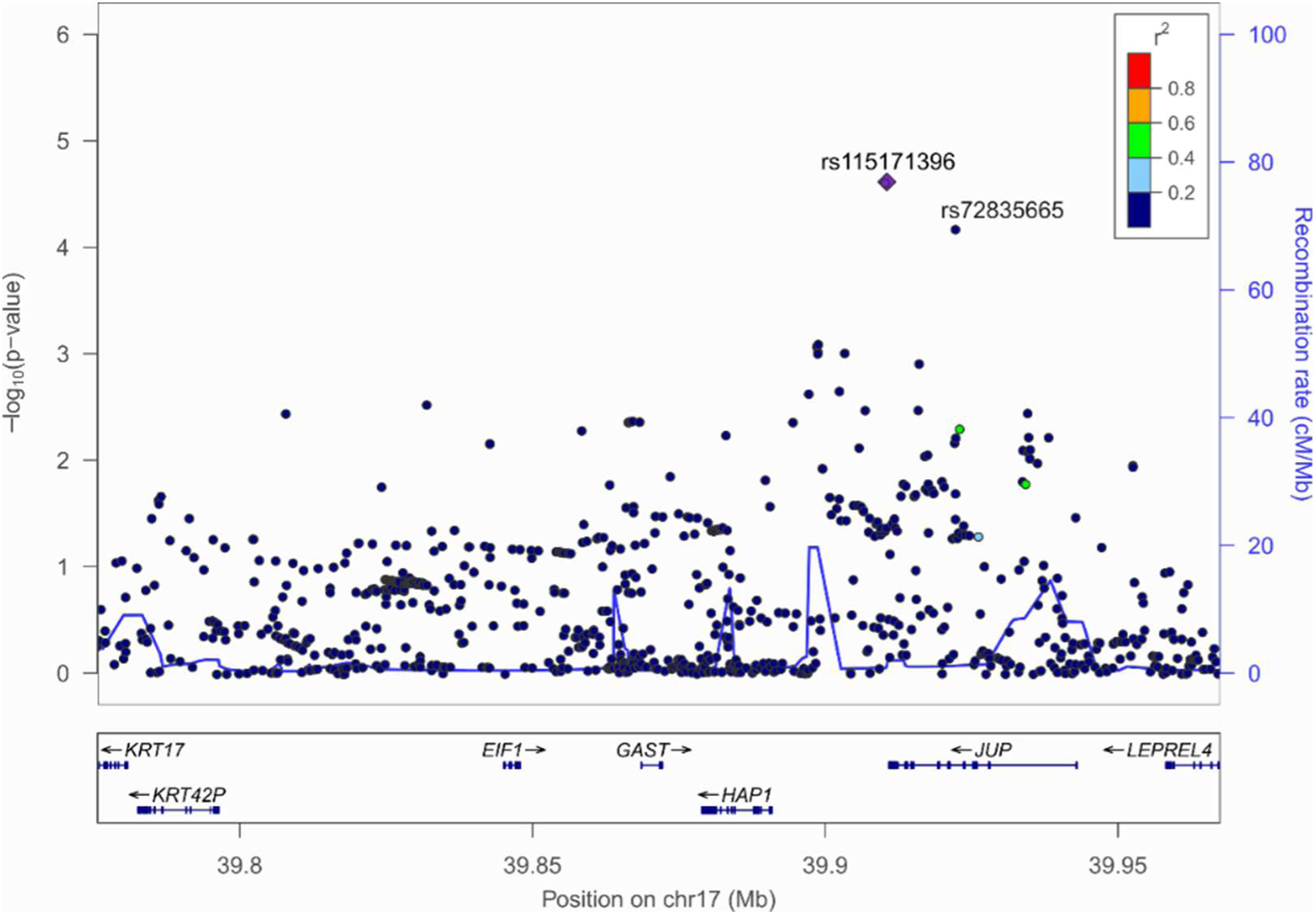
Regional association plot for P-wave over the JUP region. The purple diamond indicates the lead SNP at chr17:39910519 (rs115171396). The second, independent, significantly associated SNP rs72835665 is also displayed. Linkage disequilibrium (LD) with rs115171396 is based on the r^2^ statistic, estimated on the CHRIS sample, colored according to the legend.

**Supplementary Figure 3.**
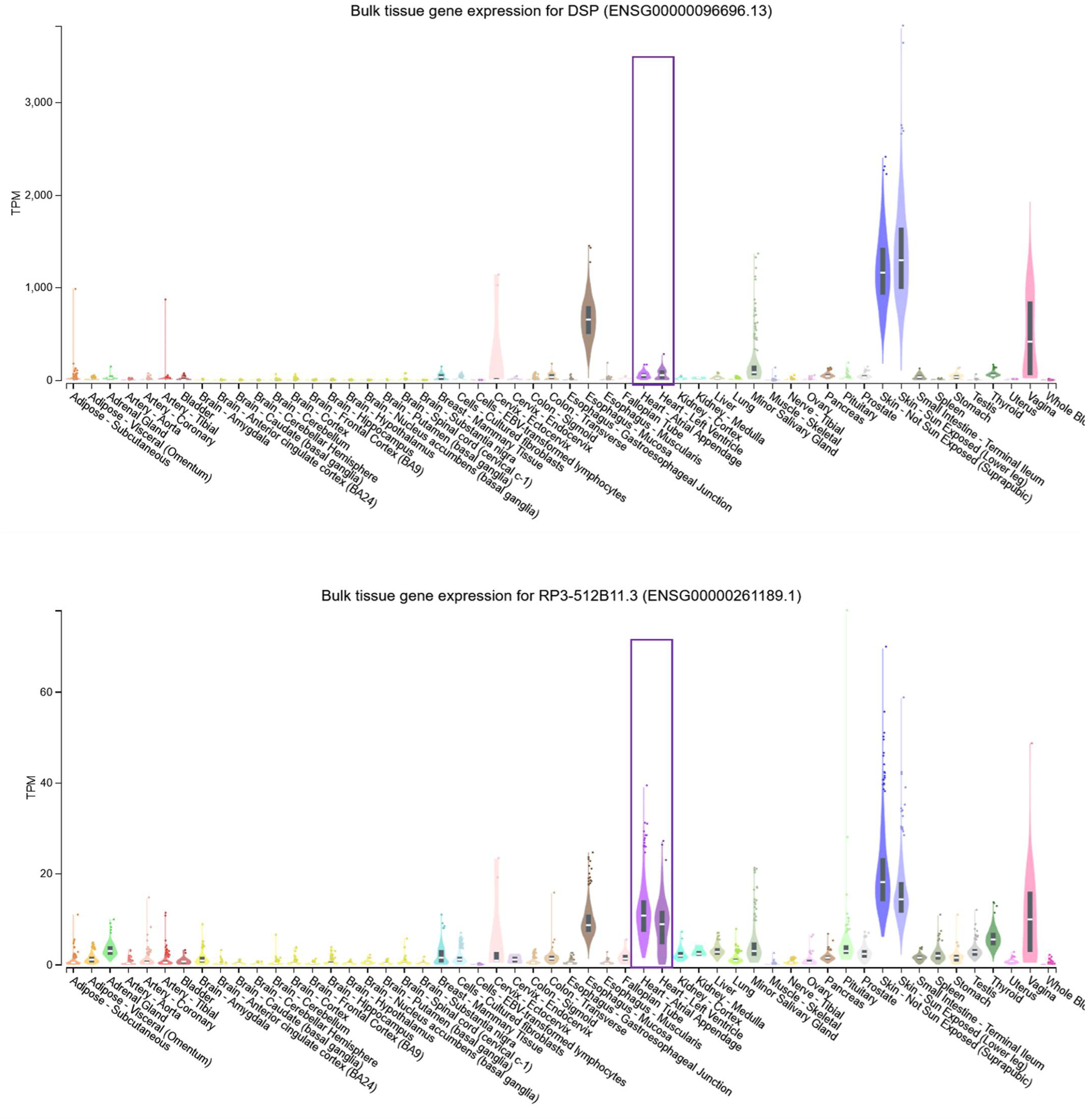
DSP and DSP-AS1 expression profile in multiple tissues. Data were downloaded from GTEx v8. *RP-3512B11.3* corresponds to *DSP-AS1*. Purple box highlights cardiac tissues available.

**Supplementary Figure 4.**
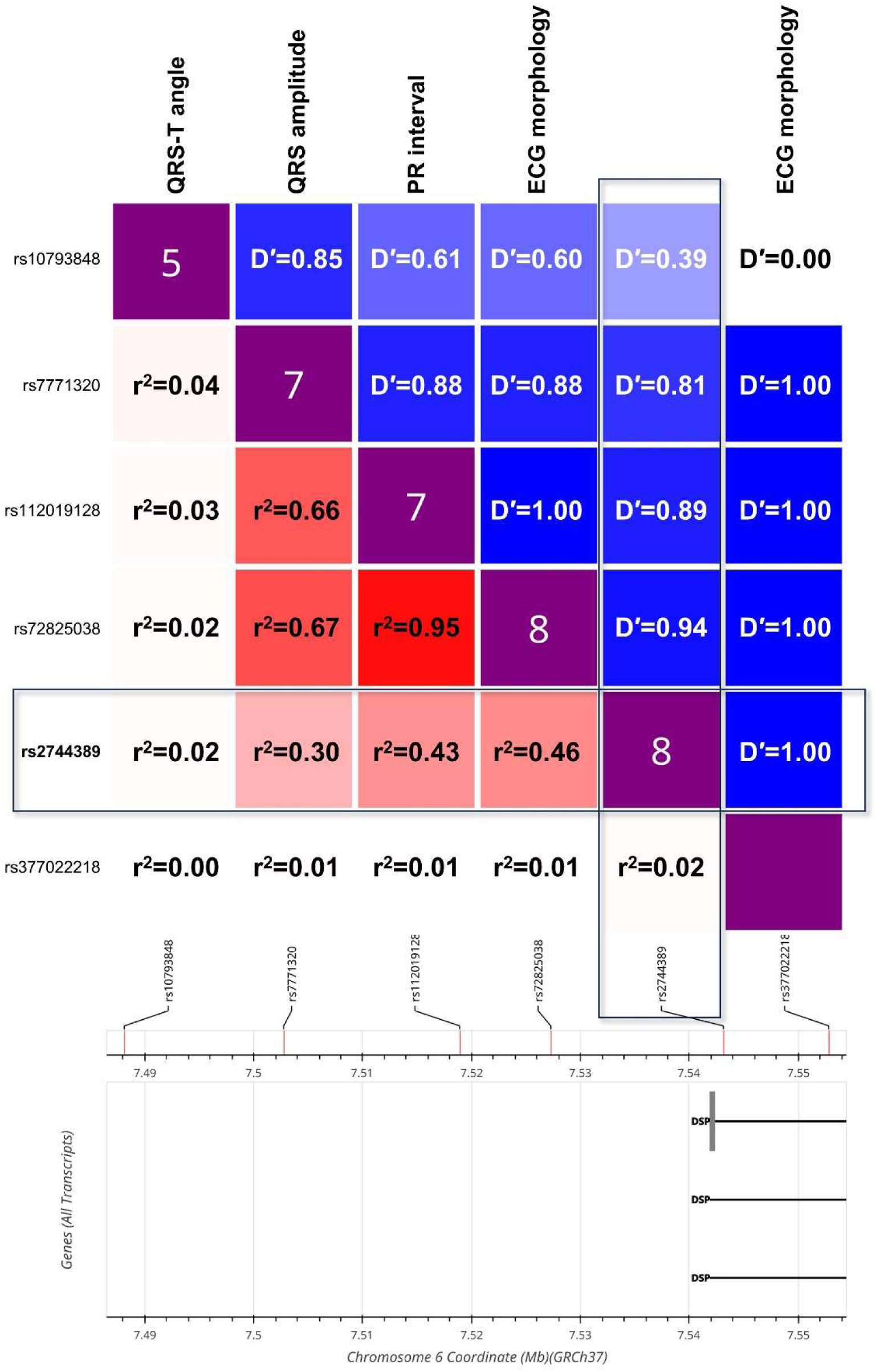
Linkage disequilibrium of rs2744389 with all nearby variants associated with ECG traits at P<5×10^-8^. rs10793848, associated with QRS-T angle (**Young 2023**); rs7771320, associated with QRS amplitude (**van der Harst 2016**); rs112019128, associated with PR interval (**Ntalla 2020**); and rs72825038 and rs377022218 associated with ECG morphology (**Verweij 2020**). The plot was generated with LDlink (https://ldlink.nih.gov/) based on 1000 Genomes CEU data. r^2^, D′, and FORGEdb scores, are reported below, above, and on, the diagonal, respectively.

**Supplementary Figure 5.**
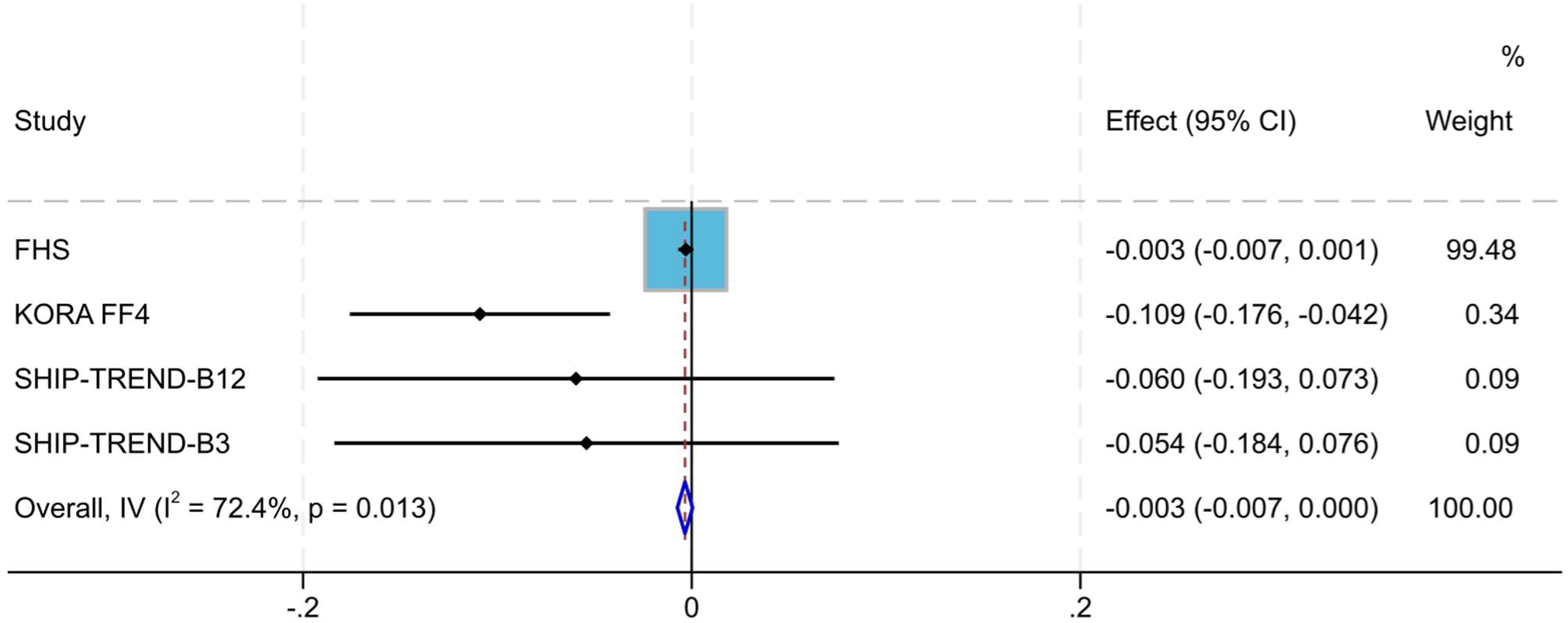
Fixed effects meta-analysis of association between rs2076298 and cg02643433. FHS, Framingham Heart Study. SHIP-TREND used two different methylation arrays, which were separately analyzed (B12 = batches 1 and 2 using EPIC array, B3 = batch 3 using EPICv2 array). The overall estimate was finally used to perform MR analysis presented in **Table 4**.

**Supplementary Figure 6.**
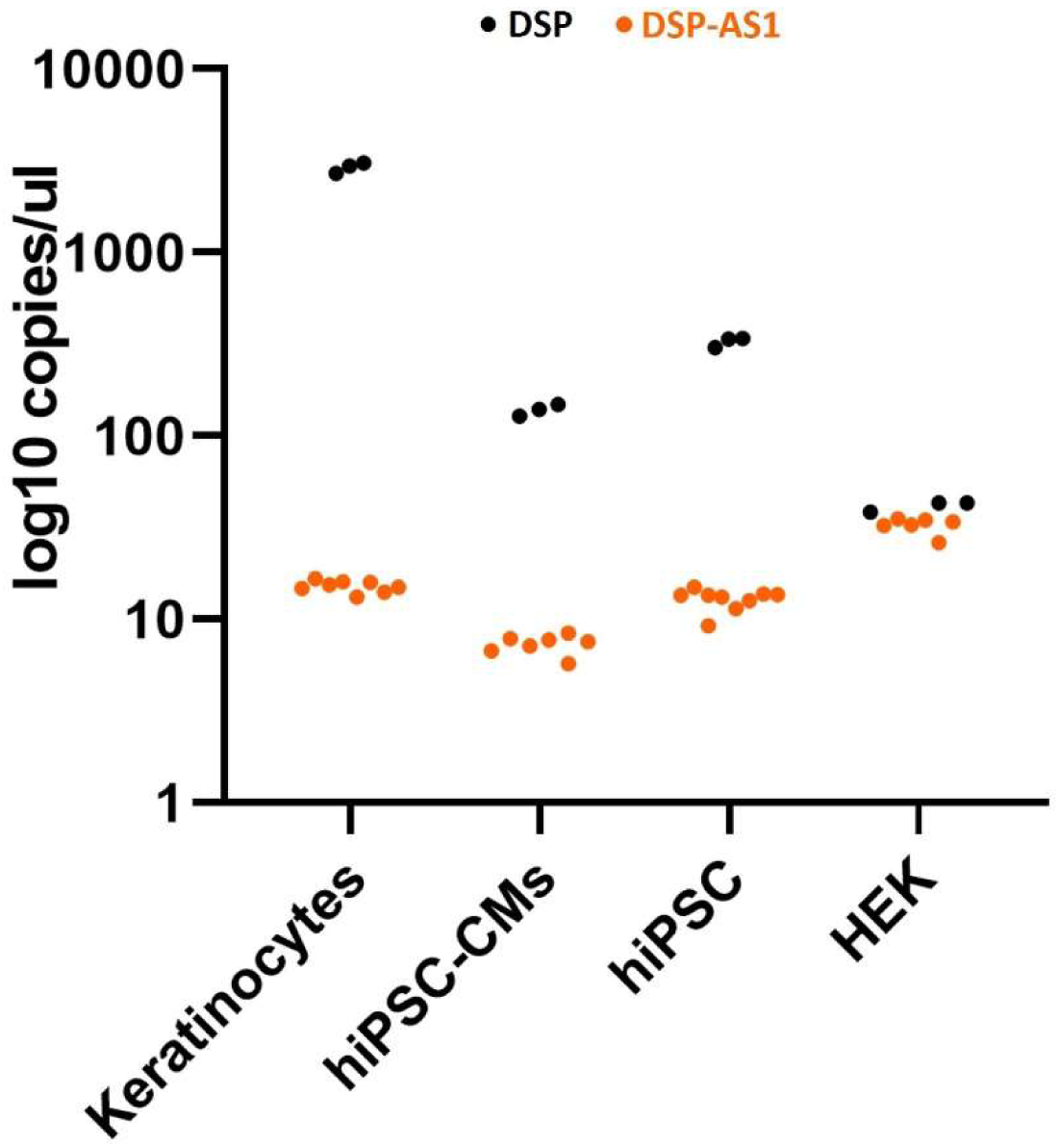
DSP-AS1 and DSP mRNA expression in different cell types. The data points in the graph correspond to biological replicates of the cell lines indicated on the x-axis, representing different passages. In the case of hiPSC-CMs, each dot represents a distinct cardiomyogenic differentiation. The raw data are reported in **Supplementary Table 11**.

**Supplementary Figure 7.**
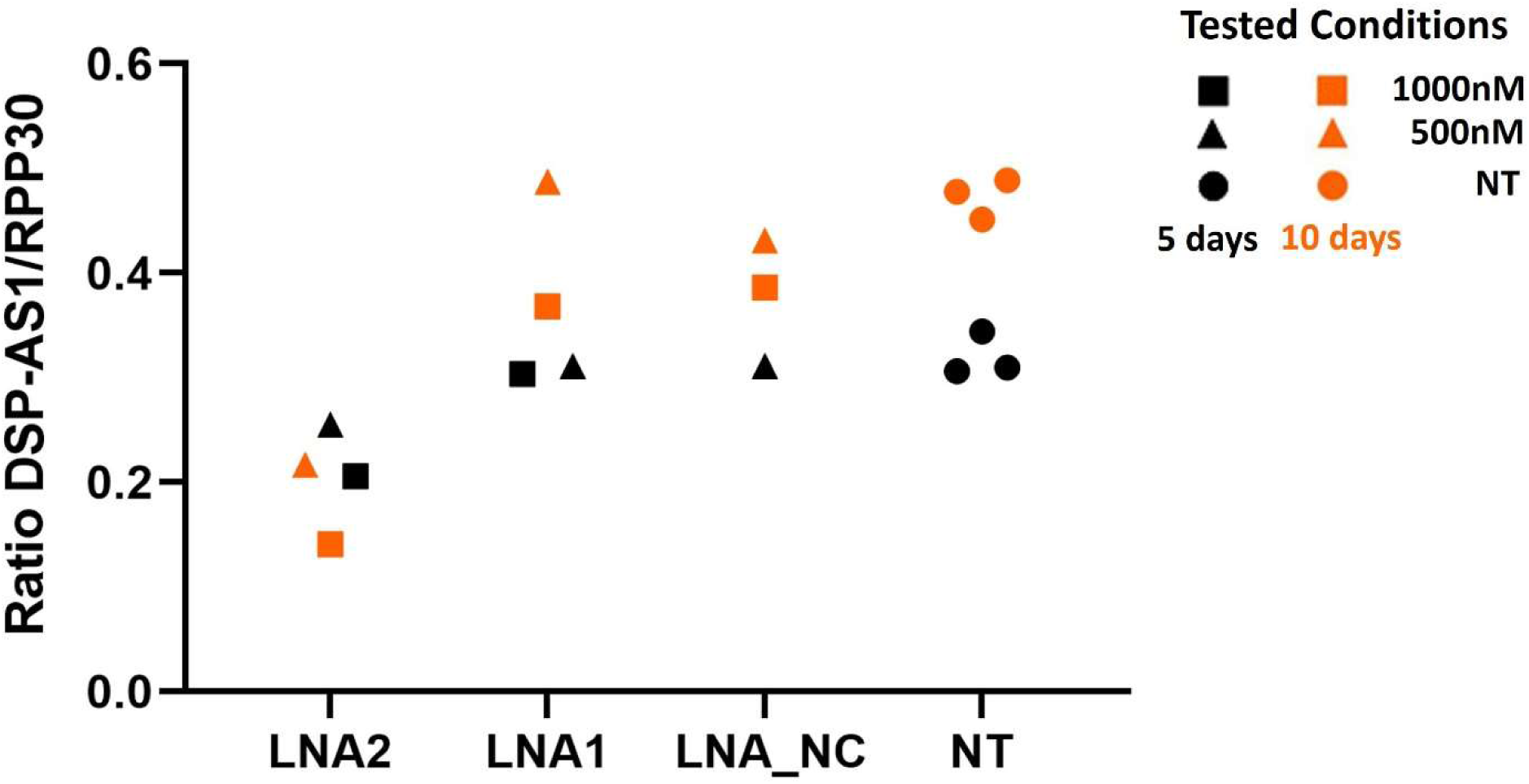
GapmeRs set up for DSP-AS1 downregulation. Two different concentrations (1000nM and 500nM) were tested for each GapmeR (LNA2, LNA1, LNA_NC). The GapmeR effect on the *DSP-AS1* expression was tested at 5 and at 10 days of treatment in hiPSC-CMs. NT: not treated cells. Two different cardiomyogenic differentiations were done. One was used for 5 days of the treatment and a second one for 10 days of treatment. Raw data are reported in **Supplementary Table 12**.

